## Supplementary File for "Serological and viral prevalence of Oropouche virus (OROV): A systematic review and meta-analysis from 2000-2024 including human, animal, and vector surveillance studies"

### Supplementary Materials

File A: PRISMA Checklist 1
File B: Search Strategy 4
File C: PROSPERO Protocol Registration 7
File D: Full inclusion and exclusion criteria 7
File E: Risk of Bias Tool Breakdown 8
Table A. Bibliographic summary of human studies. 11
Table B. Bibliographic summary of vector studies. 19
Table C. Risk of bias breakdown for all studies. 27

### File A: PRISMA Checklist

| Section and Topic | Item # | Checklist item | Location where item is reported |
|---|---|---|---|
| **TITLE** | | | |
| Title | 1 | Identify the report as a systematic review. | In title, abstract, last paragraph of introduction. |
| **ABSTRACT** | | | |
| Abstract | 2 | See the PRISMA 2020 for Abstracts checklist. | Page 1. |
| **INTRODUCTION** | | | |
| Rationale | 3 | Describe the rationale for the review in the context of existing knowledge. | Introduction, paragraph 2. Page 3. |
| Objectives | 4 | Provide an explicit statement of the objective(s) or question(s) the review addresses. | Introduction, paragraph 3. Page 3-4. |
| **METHODS** | | | |
| Eligibility criteria | 5 | Specify the inclusion and exclusion criteria for the review and how studies were grouped for the syntheses. | Methods, paragraph 2, page 4, and supplementary file D. |
| Information sources | 6 | Specify all databases, registers, websites, organisations, reference lists and other sources searched or consulted to identify studies. Specify the date when each source was last searched or consulted. | Methods, paragraph 1, page 4. |

| | | | |
|---|---|---|---|
| Search strategy | 7 | Present the full search strategies for all databases, registers and websites, including any filters and limits used. | Supplementary file B. |
| Selection process | 8 | Specify the methods used to decide whether a study met the inclusion criteria of the review, including how many reviewers screened each record and each report retrieved, whether they worked independently, and if applicable, details of automation tools used in the process. | Methods, paragraph 2, page 4. |
| Data collection process | 9 | Specify the methods used to collect data from reports, including how many reviewers collected data from each report, whether they worked independently, any processes for obtaining or confirming data from study investigators, and if applicable, details of automation tools used in the process. | Methods, paragraph 3, page 4. |
| Data items | 10a | List and define all outcomes for which data were sought. Specify whether all results that were compatible with each outcome domain in each study were sought (e.g. for all measures, time points, analyses), and if not, the methods used to decide which results to collect. | Methods, paragraph 3-5, page 4-5. Also available in the data dictionary, which is in our data availability statement and open-access database at serotracker.com , ArboTracker dashboard, also cited on the top of page 5. |
| | 10b | List and define all other variables for which data were sought (e.g. participant and intervention characteristics, funding sources). Describe any assumptions made about any missing or unclear information. | Methods, paragraph 3-5, page 4-5. Also available in the data dictionary and protocol, which are in our data availability statement and open-access database at serotracker.com , ArboTracker dashboard. |
| Study risk of bias assessment | 11 | Specify the methods used to assess risk of bias in the included studies, including details of the tool(s) used, how many reviewers assessed each study and whether they worked independently, and if applicable, details of automation tools used in the process. | Methods paragraph 4, page 5, and supplementary file E. |
| Effect measures | 12 | Specify for each outcome the effect measure(s) (e.g. risk ratio, mean difference) used in the synthesis or presentation of results. | Methods paragraph 5, page 5. |
| Synthesis methods | 13a | Describe the processes used to decide which studies were eligible for each synthesis (e.g. tabulating the study intervention characteristics and comparing against the planned groups for each synthesis (item #5)). | Methods paragraph 5, page 5. |
| | 13b | Describe any methods required to prepare the data for presentation or synthesis, such as handling of missing summary statistics, or data conversions. | Methods, paragraph 3-5, page 4-5. |
| | 13c | Describe any methods used to tabulate or visually display results of individual studies and syntheses. | Methods paragraph 5, page 5. |

| | | | |
|---|---|---|---|
| | 13d | Describe any methods used to synthesize results and provide a rationale for the choice(s). If meta-analysis was performed, describe the model(s), method(s) to identify the presence and extent of statistical heterogeneity, and software package(s) used. | Methods paragraph 5, page 5. |
| | 13e | Describe any methods used to explore possible causes of heterogeneity among study results (e.g. subgroup analysis, meta-regression). | Methods paragraph 5, page 5. |
| | 13f | Describe any sensitivity analyses conducted to assess robustness of the synthesized results. | N/A |
| Reporting bias assessment | 14 | Describe any methods used to assess risk of bias due to missing results in a synthesis (arising from reporting biases). | N/A |
| Certainty assessment | 15 | Describe any methods used to assess certainty (or confidence) in the body of evidence for an outcome. | Methods paragraph 5, page 5. |
| **RESULTS** | | | |
| Study selection | 16a | Describe the results of the search and selection process, from the number of records identified in the search to the number of studies included in the review, ideally using a flow diagram. | Results paragraph 1, page 5. Figure 1, PRISMA flowchart. |
| | 16b | Cite studies that might appear to meet the inclusion criteria, but which were excluded, and explain why they were excluded. | N/A |
| Study characteristics | 17 | Cite each included study and present its characteristics. | Page 6. |
| Risk of bias in studies | 18 | Present assessments of risk of bias for each included study. | Last paragraph of results on page 11, and supplementary file E and table C. |
| Results of individual studies | 19 | For all outcomes, present, for each study: (a) summary statistics for each group (where appropriate) and (b) an effect estimate and its precision (e.g. confidence/credible interval), ideally using structured tables or plots. | Supplementary Table A and B |
| Results of syntheses | 20a | For each synthesis, briefly summarise the characteristics and risk of bias among contributing studies. | Supplementary Table C |
| | 20b | Present results of all statistical syntheses conducted. If meta-analysis was done, present for each the summary estimate and its precision (e.g. confidence/credible interval) and measures of statistical heterogeneity. If comparing groups, describe the direction of the effect. | Figure 2-4, Results paragraph 4-6 |
| | 20c | Present results of all investigations of possible causes of heterogeneity among study results. | Discussion paragraph 6 |
| | 20d | Present results of all sensitivity analyses conducted to assess the robustness of the synthesized results. | N/A |
| Reporting biases | 21 | Present assessments of risk of bias due to missing results (arising from reporting biases) for each synthesis assessed. | N/A |
| Certainty of evidence | 22 | Present assessments of certainty (or confidence) in the body of evidence for each outcome assessed. | Figure 2-4, Results paragraph 4-6 |
| **DISCUSSION** | | | |
| Discussion | 23a | Provide a general interpretation of the results in the context of other evidence. | Human results in Paragraphs 1-4 of discussion, vector/animal |

| | | | |
|---|---|---|---|
| | | | results summarized in paragraphs 5-6. Page 11-13. |
| | 23b | Discuss any limitations of the evidence included in the review. | Discussion paragraph 7. Page 13. |
| | 23c | Discuss any limitations of the review processes used. | Discussion paragraph 7. Page 13. |
| | 23d | Discuss implications of the results for practice, policy, and future research. | Page 14. |
| **OTHER INFORMATION** | | | |
| Registration and protocol | 24a | Provide registration information for the review, including register name and registration number, or state that the review was not registered. | Abstract and methods paragraph 1. |
| | 24b | Indicate where the review protocol can be accessed, or state that a protocol was not prepared. | Methods paragraph 1 (PROSPERO) and also available at the ArboTracker online dashboard. |
| | 24c | Describe and explain any amendments to information provided at registration or in the protocol. | N/A |
| Support | 25 | Describe sources of financial or non-financial support for the review, and the role of the funders or sponsors in the review. | Conflict disclosure statement page 15 |
| Competing interests | 26 | Declare any competing interests of review authors. | Data availability statement page 15-16 |
| Availability of data, code and other materials | 27 | Report which of the following are publicly available and where they can be found: template data collection forms; data extracted from included studies; data used for all analyses; analytic code; any other materials used in the review. | Data availability statement page 15-16 |

*From:* Page MJ, McKenzie JE, Bossuyt PM, Boutron I, Hoffmann TC, Mulrow CD, et al. The PRISMA 2020 statement: an updated guideline for reporting systematic reviews. BMJ 2021;372:n71. doi: 10.1136/bmj.n71. This work is licensed under CC BY 4.0. To view a copy of this license, visit https://creativecommons.org/licenses/by/4.0/

#### File B: Search Strategy

Database: EMBASE

Dates: start date not defined to September 12, 2024

| # | **Search terms** |
|---|---|
| 1 | exp Oropouche virus/ OR exp Oropouche orthobunyavirus/ OR Oropouche.mp. OR OROV.mp. OR Iquitos virus.mp. OR IQTV.mp. OR Madre de Dios virus.mp. OR MDDV.mp. OR Perdoes virus.mp. OR PDEV.mp. |
| 2 | study.mp. OR studies.mp. OR survey*.mp. OR seroprevalence/ OR serosurvey*.mp. OR sero-survey*.mp. |

| | |
|---|---|
| | OR serosurveillance.mp. OR exp monitoring/ OR surveillance.mp. |
| 3 | exp antibody/ OR antibod*.mp. OR virus.mp. OR exp virus/ OR viral.mp. OR exp DNA/ OR DNA.mp. OR exp antigen/ OR antigen*.mp. OR seroprevalence.mp. OR exp seroprevalence/ OR sero-prevalence.mp. OR prevalence.mp. OR exp prevalence/ OR incidence.mp. OR exp incidence/ |
| 4 | detect*.mp. OR test*.mp. OR assay*.mp. OR immunoassay*.mp. OR exp immunoassay/ OR PCR.mp. OR exp polymerase chain reaction/ |
| 5 | 1 AND 2 AND 3 AND 4 |

Database: Europe PMC
Dates: January 1, 1900 to September 12, 2024
Notes: Pre-prints only

| # | Search terms |
|---|---|
| 1 | Oropouche OR "ORO virus" OR oropuche OR ORVO OR "Madre de Dios virus" OR MDDV OR "Perdoes virus" OR PDEV OR "Iquitos virus" OR IQTV |
| 2 | study OR studies OR survey* OR serosurvey* OR sero-survey* OR surveillance |
| 3 | antibod* OR virus OR viral OR DNA OR antigen OR seroprevalence OR sero-prevalence OR prevalence OR incidence |
| 4 | detect* OR test* OR assay* OR immunoassay* OR PCR |
| 5 | 1 AND 2 AND 3 AND 4 |
| 6 | AND (FIRST_PDATE:[1900-01-01 TO 2024-09-12]) |
| 7 | AND (SRC:PPR) |

Database: LILACS
Dates: start date not defined to September 12, 2024

| # | Search terms |
|---|---|
| 1 | oropouche OR OROV OR "Iquitos virus" OR IQTV OR "Madre de Dios virus" OR MDDV OR "Perdoes virus" OR PDEV |
| 2 | study OR studies OR survey* OR serosurvey OR sero-survey* OR surveillance OR sero-surveillance OR serosurveillance |
| 3 | detect* OR test* OR assay* OR immunoassay* OR PCR |
| 4 | antibod* OR virus OR viral OR DNA OR antigen* OR seroprevalence OR sero-prevalence OR prevalence OR incidence |
| 5 | 1 AND 2 AND 3 AND 4 |

Database: Medline

Dates: start date not defined to September 12, 2025

| # | Search terms |
|---|---|
| 1 | exp Oropouche virus/ OR exp Oropouche orthobunyavirus/ OR Oropouche.mp. OR OROV.mp. |
| 2 | study.mp. OR studies.mp. OR survey*.mp. OR serosurvey*.mp. OR sero-survey*.mp. OR serosurveillance.mp. OR exp monitoring/ OR surveillance.mp. |
| 3 | exp antibody/ OR antibod*.mp. OR virus.mp. OR exp virus/ OR viral.mp. OR exp DNA/ OR DNA.mp. OR exp antigen/ OR antigen*.mp. OR seroprevalence.mp. OR exp seroprevalence/ OR sero-prevalence.mp. OR prevalence.mp. OR exp prevalence/ OR incidence.mp. OR exp incidence/ |
| 4 | detect*.mp. OR test*.mp. OR assay*.mp. OR immunoassay*.mp. OR exp immunoassay/ OR PCR.mp. OR exp polymerase chain reaction/ |
| 5 | 1 AND 2 AND 3 AND 4 |

Database: Pubmed
Dates: start date not defined to September 12, 2024

| # | Search terms |
|---|---|
| 1 | "oropouche"[All Fields] OR "OROV"[All Fields] OR "Iquitos virus" [All Fields] OR "IQTV" [All Fields] OR "Madre de Dios virus" [All Fields] OR "MDDV" [All Fields] OR "Perdoes virus" [All Fields] OR "PDEV" [All Fields] |
| 2 | "studies"[All Fields] OR "study"[All Fields] OR "study s"[All Fields] OR "studying"[All Fields] OR "studys"[All Fields] OR "studies"[All Fields] OR "study"[All Fields] OR "study s"[All Fields] OR "studying"[All Fields] OR "studys"[All Fields] OR "survey*"[All Fields] OR "serosurvey*"[All Fields] OR "sero survey*"[All Fields] OR "serosurveillance"[All Fields] OR "epidemiology"[MeSH Subheading] OR "epidemiology"[All Fields] OR "surveillance"[All Fields] OR "epidemiology"[MeSH Terms] OR "surveilance"[All Fields] OR "surveillances"[All Fields] OR "surveilled"[All Fields] OR "surveillence"[All Fields] |
| 3 | "antibod*"[All Fields] OR ("virology"[MeSH Subheading] OR "virology"[All Fields] OR "viruses"[All Fields] OR "viruses"[MeSH Terms] OR "virus s"[All Fields] OR "viruse"[All Fields] OR "virus"[All Fields]) OR ("virally"[All Fields] OR "virals"[All Fields] OR "virology"[MeSH Terms] OR "virology"[All Fields] OR "viral"[All Fields]) OR ("dna"[MeSH Terms] OR "dna"[All Fields]) OR "antigen*"[All Fields] OR ("seroepidemiologic studies"[MeSH Terms] OR ("seroepidemiologic"[All Fields] AND "studies"[All Fields]) OR "seroepidemiologic studies"[All Fields] OR "seroprevalence"[All Fields] OR "seroprevalences"[All Fields] OR "seroprevalance"[All Fields] OR "seroprevalances"[All Fields] OR "seroprevalency"[All Fields] OR "seroprevalent"[All Fields]) OR "sero-prevalence"[All Fields] OR ("epidemiology"[MeSH Subheading] OR "epidemiology"[All Fields] OR "prevalence"[All Fields] OR "prevalence"[MeSH Terms] OR "prevalance"[All Fields] OR "prevalences"[All Fields] OR "prevalence s"[All Fields] OR "prevalent"[All Fields] OR "prevalently"[All Fields] OR "prevalents"[All Fields]) OR ("epidemiology"[MeSH Subheading] OR "epidemiology"[All Fields] OR "incidence"[All Fields] OR "incidence"[MeSH Terms] OR "incidences"[All Fields] OR "incident"[All Fields] OR "incidents"[All Fields]) |
| 4 | "detect*"[All Fields] OR "test*"[All Fields] OR "assay*"[All Fields] OR "immunoassay*"[All Fields] OR "PCR"[All Fields] |
| 5 | 1 AND 2 AND 3 AND 4 |

Database: Web of Science
Dates: start date not defined to September 12, 2024

| # | Search terms |
|---|---|
| 1 | ALL=(oropouche OR ORVO OR "Madre de Dios virus" OR MDDV OR "Perdoes virus" OR PDEV OR "Iquitos virus" OR IQTV) |
| 2 | ALL = (study OR studies OR survey* OR serosurvey* OR sero-survey* OR surveillance) |
| 3 | ALL=(antibod* OR virus OR viral OR DNA OR antigen OR seroprevalence OR sero-prevalence OR prevalence OR incidence) |
| 4 | ALL=(detect* OR test* OR assay* OR immunoassay* OR PCR) |
| 5 | 1 AND 2 AND 3 AND 4 |

#### File C: PROSPERO Protocol Registration

The following protocol is pulled from our most recent PROSPERO protocol registration (March 17, 2025 version) CRD42024551000.

https://www.crd.york.ac.uk/PROSPERO/view/CRD42024551000

#### File D: Full inclusion and exclusion criteria

Criteria for including evidence (must meet all the criteria to be included)

| Characteristics | Criteria for inclusion |
|---|---|
| Population | ● Humans of any age<br>● Animal populations<br>● Insect populations<br>● Including studies that only included symptomatic individuals or those with suspected OROV |
| Study design | ● Sero-surveys – defined as the collection and testing of serum (or proxy such as oral fluid) specimens to estimate the prevalence of antibodies or T-cells against OROV as an indicator of immunity, and/or<br>● Molecular or viral epidemiology studies to estimate the positivity rate of direct virus detection (through any of the following: PCR, sequencing, immunofluorescence, antigen) for OROV<br>● Samples from a defined population over a specified period of time, including symptomatic persons and those with suspected disease<br>● Cross-sectional, repeated cross-sectional, evaluations of serological tests, case-control, and cohort study designs, with serology or PCR measurements at single time points or repeated at multiple time points |
| *Special design | ● Include systematic reviews and meta-analyses of seroprevalence studies for the purpose of tracking evidence-synthesis efforts |
| Sampling | ● Any sampling method |
| Types of evidence | ● Published or preprinted academic literature, |

| | |
|---|---|
| | • Grey literature (government, institutional, or meeting reports)<br>• Media reports<br>• Slide deck presentations were included if we could identify the person giving the presentation and the date of the presentation, and the institution |
| Outcome measures | • Reports a seroprevalence estimate (proportion of the population with detectable antibodies; including negative results i.e. undetectable antibodies), or<br>• Reports a prevalence estimate (proportion of the population with detectable viral nucleic acids; including negative results i.e. undetectable nucleic acids)<br>• Reports the number of participants enrolled in the study (denominator)<br>• Reports study sampling period (date or week, can be inferred from month/year)<br>• Reports the locations at which the study took places such that they could be categorized as neighbourhood, city, state/province/territory, or country |
| Languages | • Any |

Criteria for excluding evidence (if any met then exclude)

| Characteristics | Criteria for exclusion |
|---|---|
| Population | • Laboratory/experimental (non-field study population: e.g., *in silico*, *in vitro;* non-natural infection) |
| Study design | • Study designs other than cross-sectional or cohort design (such as case reports, study protocols) |
| Sampling | • N/A |
| Types of evidence | • Multimedia sources of data (audio clips, video clips) were excluded due to the feasibility of extracting. Slide deck presentations were excluded if we could not identify the person giving the presentation and the date of the presentation |
| Outcome measures | • Does not report OROV prevalence or sufficient information to calculate a prevalence of OROV<br>• Does not report study sampling end date/week, or cannot be reasonably inferred with the available information<br>• Does not report the number of participants included in the study (sample denominator)<br>• Does not report the location at which the study took place |
| Language | • N/A |

#### File E: Risk of Bias Tool Breakdown

To assess risk of bias, a decision rule assigned a rating of low, moderate, or high risk of bias to each study based on the specific combination of JBI checklist ratings for that study [1]. This decision rule was developed based on guidance on estimating disease prevalence [17,18] and was validated against assessments derived manually by two independent reviewers for 2,070 seroprevalence studies in the SeroTracker database, showing good agreement with manual review (intraclass correlation 0.77, 95% CI 0.74-0.80) in a recent paper [1].

| **Item 1: Was the sample frame appropriate to address the target population?** | |
|---|---|
| Yes | Sample frame described and approximated the target population |
| No | Sample frame did not approximate the target population (e.g., blood donors do not represent general population, doctors do not represent all health care providers) |
| Exclude | Sample frame not described |
| *Notes | The term “target population” should not be taken to infer every individual from everywhere or with similar disease or exposure characteristics. Instead, give consideration to specific population characteristics in the study, including age range, gender, morbidities, medications, and other potentially influential factors. For example, a sample frame may not be appropriate to address the target population if a certain group has been used (such as those working for one organisation, or one profession) and the results then inferred to the target population (i.e. working adults). A sample frame may be appropriate when it includes almost all the members of the target population (i.e. a census, or a complete list of participants or complete registry data). |

| **Item 2: Were study participants recruited in an appropriate way?** | |
|---|---|
| Yes | Convenience sampling, probability sampling method (simple or stratified random) or entire sample (e.g., an entire town) was used |
| No | Sampling method not reported |
| Exclude | |

| **Item 3: Was the sample size adequate?** | |
|---|---|
| Yes | ≥99 |
| No | <99 |
| Exclude | Sample size not reported |
| *Notes | |

| **Item 4: Were the study subjects and setting described in detail?** | |
|---|---|
| Yes | Average age and distribution of gender/sex provided |
| No | Neither age or gender/sex is provided, or only one of age and gender/sex is provided |

| Item 5: Was data analysis conducted with sufficient coverage of the identified sample? | |
|---|---|
| Yes | The demographic characteristics (gender/sex, age, and ethnicity) of the sample are at least somewhat representative of the population in both the main and sub-group analyses |
| No | The demographic characteristics (gender/sex, age, and ethnicity) of the sample are not representative of the population in both the main and sub-group analyses |
| Unclear | Information is not provided about demographic characteristics of the sample (gender/sex, age, and ethnicity) |

| Item 6: Were valid methods used for the identification of the condition? | |
|---|---|
| Yes | Serology or viral test type was reported |
| No | Serology or viral test type was not reported |
| Exclude | |

| Item 7: Was the condition measured in a standard, reliable way for all participants? | |
|---|---|
| Yes | The same serology test was used for all participants |
| No | Different serology tests were used for participants |
| Unclear | No details were provided about which participants received which serology tests |

| Item 8: Was there appropriate statistical analysis? | |
|---|---|
| Yes | Corrects for population characteristics OR the sample is somewhat representative of the population, and provides the information necessary to determine the numerator, denominator, prevalence estimate, and confidence interval. |
| No | Does not correct for population characteristics and the sample is not likely representative of the population or does not provide the information necessary to determine the numerator, denominator, prevalence estimate, and confidence interval. |

| Item 9: Overall risk of bias | |
|---|---|
| Low | The estimates are very likely correct for the target population. To obtain a low risk of bias classification, all criteria must be met or departures from the criteria must be minimal and unlikely to impact on the validity and reliability of the prevalence estimate. These include sample sizes that are just below the threshold when all other criteria are met, reporting only some of characteristics of the sample, test characteristics below the threshold but corrections for the test performance, and response rates that are just below the threshold in the context of probability based sampling of an appropriate sampling frame with population weighted seroprevalence estimates. |
| Moderate | The estimates are likely correct for the target population. To obtain a moderate risk of bias classification, most criteria must be met and departures from the criteria are likely to have only a small impact on the validity and reliability of the prevalence estimates. |

| | |
|---|---|
| High | The estimates are not likely correct for the target population. To obtain a high risk of bias, many criteria must not be met or departures from criteria are likely to have a major impact on the validity and reliability of the prevalence estimates. |
| Unclear | There was insufficient information to assess the risk of bias. |


### Table A. Bibliographic summary of human studies.

| Author | Sampling year | Country | Population | Estimate type | Positive cases | Number tested | Prevalence | Assay type | Assay target |
|---|---|---|---|---|---|---|---|---|---|
| F. P. Pinheiro[1] | 1975 | Brazil | Students and Daycares | Seroprevalence | 45 | 112 | 0.402 | HAI | NAb |
| James LeDuc[2] | 1978 | Brazil | Febrile patients | Seroprevalence | 164 | 555 | 0.295 | HAI | NR |
| Ronaldo B. Freitas[3] | 1979 | Brazil | Febrile patients | Viral Prevalence | 57 | 546 | 0.104 | Viral isolation | NAb |
| Ronaldo B. Freitas[3] | 1979 | Brazil | Community | Seroprevalence | 381 | 2,975 | 0.128 | HAI | NAb |
| Carlos Borborema[4] | 1980 | Brazil | Community | Seroprevalence | 110 | 1,018 | 0.108 | HAI | NAb |
| Pedro Fernando da Costa Vasconcelos[5] | 1988 | Brazil | Febrile patients | Seroprevalence | 256 | 394 | 0.650 | Other | IgM,NAb |
| Douglas Watts[6] | 1994 | Peru | Essential non-healthcare workers | Seroprevalence | 6 | 68 | 0.088 | ELISA | IgM |
| Amélia Rosa[7] | 1994 | Brazil | Community | Seroprevalence | 490 | 592 | 0.828 | Other | IgM,NAb |
| Kathy Baisley[8] | 1996 | Peru | Community | Seroprevalence | 828 | 2,454 | 0.337 | ELISA | IgG |
| José Tavares-Neto[9] | 1999 | Brazil | Community | Seroprevalence | 20 | 380 | 0.053 | Other | NR |
| Douglas M. Watts[10] | 1999 | Peru | Febrile patients | Seroprevalence | 203 | 19,798 | 0.010 | ELISA | IgG,IgM |
| Regina Maria Pinto De Figueiredo[11] | 1999 | Brazil | Residual sera | Seroprevalence | 3 | 35 | 0.086 | ELISA | IgM |
| Raimunda do Socorro da Silva Azevedo[12] | 2004 | Brazil | Positive (PCR) or suspected cases | Seroprevalence | 284 | 734 | 0.387 | ELISA | IgM |
| Stephen Manock[13] | 2004 | Ecuador | Febrile patients | Seroprevalence | 2 | 608 | 0.003 | ELISA | IgM |

| | | | | | | | | | |
|---|---|---|---|---|---|---|---|---|---|
| Marcio Nunes[14] | 2005 | Brazil | Other | Seroprevalence | 22 | 2,766 | 0.008 | HAI | NAb |
| Helena Vasconcelos[15] | 2006 | Brazil | Febrile patients | Seroprevalence | 480 | 1,113 | 0.431 | HAI | NAb |
| Ana Carolina Bernardes Terzian[16] | 2006 | Brazil | Febrile patients | Viral Prevalence | 1 | 69 | 0.014 | RT-PCR | S segment (OROV only) |
| Patricia Aguilar[17] | 2006 | Peru | Febrile patients | Seroprevalence | 154 | 1,037 | 0.149 | ELISA | IgG |
| Brett Forshey[18] | 2007 | Bolivia (Plurinational State of) | Febrile patients | Viral Prevalence | 593 | 42,354 | 0.014 | RT-PCR | NR |
| Ana Cecilia Ribeiro Cruz[19] | 2008 | Brazil | Community | Seroprevalence | 113 | 3,194 | 0.035 | HAI | NAb |
| Maria Paula Mourão[20] | 2008 | Brazil | Febrile patients | Seroprevalence | 128 | 631 | 0.203 | ELISA | IgM |
| Pedro P. Alvarez[21] | 2010 | Peru | Febrile patients | Seroprevalence | 108 | 675 | 0.160 | ELISA | IgM |
| Valquiria do Carmo Alves Martins[22] | 2011 | Brazil | Positive cases of a different arbovirus | Viral Prevalence | 0 | 677 | 0.000 | RT-PCR | NR |
| Sara Castro[23] | 2011 | Peru | Positive (PCR) or suspected cases | Seroprevalence | 17 | 26 | 0.654 | ELISA | IgM |
| Sara Castro[23] | 2011 | Peru | Positive (PCR) or suspected cases | Viral Prevalence | 0 | 26 | 0.000 | RT-PCR | NR |
| Belgath Fernandes Cardoso[24] | 2012 | Brazil | Febrile patients | Viral Prevalence | 5 | 524 | 0.010 | RT-PCR | S segment (OROV only) |
| Michele S. Bastos[25] | 2012 | Brazil | Non-arboviral patients | Viral Prevalence | 6 | 330 | 0.018 | RT-PCR | S segment (OROV only) |
| Vivaldo Gomes da Costa[26] | 2013 | Brazil | Positive (PCR) or suspected cases | Seroprevalence | 10 | 1,294 | 0.008 | ELISA | IgM |
| Maha Elbadry[27] | 2014 | Haiti | Febrile patients | Viral Prevalence | 1 | 1,250 | 0.001 | RT-PCR | NR |
| Raquel Curtinhas de Lima[28] | 2015 | Brazil | Febrile patients | Seroprevalence | 17 | 166 | 0.102 | PRNT | NAb |
| Raquel Curtinhas de Lima[28] | 2015 | Brazil | Febrile patients | Viral Prevalence | 0 | 166 | 0.000 | RT-PCR | S segment (OROV only) |
| Felipe Naveca[29] | 2015 | Brazil | Febrile patients | Viral Prevalence | 9 | 30 | 0.300 | RT-PCR | S segment (OROV only) |

| | | | | | | | | | |
|---|---|---|---|---|---|---|---|---|---|
| Barbara Batista Salgado[30] | 2015 | Brazil | Essential non-healthcare workers | Seroprevalence | 4 | 595 | 0.007 | HAI | NAb |
| Maria Garcia[31] | 2016 | Peru | Febrile patients | Seroprevalence | 122 | 508 | 0.240 | ELISA | IgM |
| Maria Garcia[31] | 2016 | Peru | Febrile patients | Viral Prevalence | 51 | 1,016 | 0.050 | RT-PCR | NR |
| Tung Gia Phan[32] | 2016 | Peru | Febrile patients | Viral Prevalence | 2 | 18 | 0.111 | RT-PCR | NR |
| Carlos Alva-Urcia[33] | 2016 | Peru | Febrile patients | Viral Prevalence | 24 | 278 | 0.086 | RT-PCR | NR |
| Helver Dias[34] | 2016 | Brazil | Febrile patients | Viral Prevalence | 0 | 106 | 0.000 | RT-PCR | S segment (OROV only) |
| Valdinete Alves do Nascimento[35] | 2016 | Brazil | Positive (PCR) or suspected cases | Viral Prevalence | 5 | 352 | 0.014 | RT-PCR | NR |
| Juana del Valle-Mendoza[36] | 2016 | Peru | Febrile patients | Viral Prevalence | 0 | 248 | 0.000 | RT-PCR | NR |
| Hilda Durango-Chavez[37] | 2016 | Peru | Febrile patients | Viral Prevalence | 151 | 741 | 0.204 | RT-PCR | NR |
| M.C. de Souza Costa[38] | 2016 | Brazil | Febrile patients | Viral Prevalence | 40 | 897 | 0.045 | RT-PCR | S segment (OROV only) |
| Wilmer Silva-Caso[39] | 2016 | Peru | Febrile patients | Viral Prevalence | 92 | 536 | 0.172 | RT-PCR | S segment (OROV only) |
| Wilmer Silva-Caso[40] | 2016 | Peru | Febrile patients | Viral Prevalence | 46 | 268 | 0.172 | RT-PCR | NR |
| Johanna Martins-Luna[41] | 2016 | Peru | Febrile patients | Viral Prevalence | 131 | 496 | 0.264 | RT-PCR | NR |
| Johanna Martins-Luna[42] | 2016 | Peru | Febrile patients | Viral Prevalence | 393 | 1,488 | 0.264 | RT-PCR | NR |
| Juana del Valle-Mendoza[43] | 2016 | Peru | Febrile patients | Viral Prevalence | 0 | 95 | 0.000 | RT-PCR | NR |
| Cassiano Junior Saatkamp[44] | 2016 | Brazil | Febrile patients | Viral Prevalence | 0 | 49 | 0.000 | RT-PCR | S segment (OROV only) |
| Emma L. Wise[45] | 2016 | Ecuador | Febrile patients | Viral Prevalence | 6 | 258 | 0.023 | RT-PCR | S segment (OROV only) |
| Marco Coaguila[46] | 2017 | Peru | Febrile patients | Seroprevalence | 0 | 1,983 | 0.000 | Other | NR |
| Juliana Gil-Mora[47] | 2017 | Colombia | Community | Seroprevalence | 10 | 505 | 0.020 | PRNT | NAb |

| | | | | | | | | | |
|---|---|---|---|---|---|---|---|---|---|
| Larissa Moraes dos Santos Fonseca[48] | 2017 | Brazil | Febrile patients | Viral Prevalence | 5 | 53 | 0.094 | RT-PCR | S segment (OROV only) |
| Vanessa L. Carvalho[49] | 2018 | Brazil | Febrile patients | Seroprevalence | 37 | 90 | 0.411 | ELISA | IgM |
| Janeth Aracely Ramirez Pavon[50] | 2019 | Brazil | Positive (PCR) or suspected cases | Viral Prevalence | 1 | 6 | 0.167 | RT-PCR | S segment (OROV only) |
| Luiz Henrique Gonçalves Maciel[51] | 2019 | Brazil | Non-arboviral patients | Viral Prevalence | 0 | 340 | 0.000 | RT-PCR | S segment (OROV only) |
| Jackson Alves da Silva Queiroz[52] | 2019 | Brazil | Febrile patients | Viral Prevalence | 0 | 308 | 0.000 | RT-PCR | NR |
| Diego Michel Fernandes da Silva[53] | 2020 | Brazil | Positive (PCR) or suspected cases | Viral Prevalence | 0 | 79 | 0.000 | RT-PCR | NR |
| Mélanie Gaillet[54] | 2020 | French Guiana | Febrile patients | Viral Prevalence | 23 | 28 | 0.821 | RT-PCR | NR |
| Carlos Silva-Ramos[55] | 2021 | Colombia | Febrile patients | Viral Prevalence | 0 | 116 | 0.000 | RT-PCR | NR |
| Karl Ciuoderis[56] | 2022 | Colombia | Febrile patients | Seroprevalence | 234 | 1,652 | 0.142 | MIA | IgG |
| Karl Ciuoderis[56] | 2022 | Colombia | Febrile patients | Viral Prevalence | 174 | 1,582 | 0.110 | RT-PCR | M segment (OROV only) |
| Hillquias Monteiro Moreira[57] | 2023 | Brazil | Febrile patients | Viral Prevalence | 27 | 351 | 0.077 | RT-PCR | S segment (OROV only),M segment (OROV only) |
| Liliana Sanchez-Lerma[58] | 2023 | Colombia | Febrile patients | Viral Prevalence | 0 | 100 | 0.000 | RT-PCR | N Segment |
| Gabriel Scachetti[59] | 2024 | Brazil | Febrile patients | Viral Prevalence | 10 | 93 | 0.108 | RT-PCR | NR |

#### Table B. Bibliographic summary of vector studies.

| Author | Sampling year | Country | Population | Species | Estimate type | Positive cases | Number tested | Prevalence | Assay type | Assay target |
|---|---|---|---|---|---|---|---|---|---|---|
| F. P. Pinheiro[1] | 1975 | Brazil | Insect | Culicoides paraensis | Viral Prevalence | 2 | 15,000 | 0.000 | Viral isolation | NAb |
| F. P. Pinheiro[1] | 1975 | Brazil | Non-human animal | Multiple | Seroprevalence | 47 | 1,494 | 0.031 | HAI | NAb |
| F. P. Pinheiro[1] | 1975 | Brazil | Non-human animal | Multiple | Seroprevalence | 47 | 1,494 | 0.031 | HAI | NAb |
| F. P. Pinheiro[1] | 1975 | Brazil | Non-human animal | Multiple | Seroprevalence | 47 | 1,494 | 0.031 | HAI | NAb |
| F. P. Pinheiro[1] | 1975 | Brazil | Non-human animal | Multiple | Seroprevalence | 47 | 1,494 | 0.031 | HAI | NAb |
| Carlos Borborema[2] | 1980 | Brazil | Insect | Culex quinquefasciatus | Seroprevalence | 1 | 42 | 0.024 | HAI | NAb |
| Carlos Borborema[2] | 1980 | Brazil | Insect | Culex quinquefasciatus | Seroprevalence | 1 | 42 | 0.024 | HAI | NAb |
| Carlos Borborema[2] | 1980 | Brazil | Insect | Culex quinquefasciatus | Seroprevalence | 1 | 42 | 0.024 | HAI | NAb |
| Carlos Borborema[2] | 1980 | Brazil | Insect | Culicoides paraensis | Seroprevalence | 0 | 147 | 0.000 | HAI | NAb |
| Carlos Borborema[2] | 1980 | Brazil | Insect | Culicoides paraensis | Seroprevalence | 0 | 147 | 0.000 | HAI | NAb |

| | | | | | | | | | | |
|---|---|---|---|---|---|---|---|---|---|---|
| PF da Costa Vasconcelos[3] | 1988 | Brazil | Insect | Aedes scapularis | Seroprevalence | 0 | 1 | 0.000 | CF | IgM,NAb |
| PF da Costa Vasconcelos[3] | 1988 | Brazil | Insect | Anopheles nuneztovari | Seroprevalence | 0 | 1 | 0.000 | CF | IgM,NAb |
| PF da Costa Vasconcelos[3] | 1988 | Brazil | Insect | Anopheles triannulatus | Seroprevalence | 0 | 1 | 0.000 | CF | IgM,NAb |
| PF da Costa Vasconcelos[3] | 1988 | Brazil | Insect | Culex Carrolia Sp | Seroprevalence | 0 | 1 | 0.000 | CF | IgM,NAb |
| PF da Costa Vasconcelos[3] | 1988 | Brazil | Insect | Culex Spp. | Seroprevalence | 0 | 1 | 0.000 | CF | IgM,NAb |
| PF da Costa Vasconcelos[3] | 1988 | Brazil | Insect | Culex corniger | Seroprevalence | 0 | 1 | 0.000 | CF | IgM,NAb |
| PF da Costa Vasconcelos[3] | 1988 | Brazil | Insect | Culex coronator | Seroprevalence | 0 | 1 | 0.000 | CF | IgM,NAb |
| PF da Costa Vasconcelos[3] | 1988 | Brazil | Insect | Culex declarator | Seroprevalence | 0 | 3 | 0.000 | CF | IgM,NAb |
| PF da Costa Vasconcelos[3] | 1988 | Brazil | Insect | Culex quinquefasciatus | Seroprevalence | 0 | 79 | 0.000 | CF | IgM,NAb |
| PF da Costa Vasconcelos[3] | 1988 | Brazil | Insect | Culicoides paraensis | Seroprevalence | 1 | 39 | 0.026 | CF | IgM,NAb |
| PF da Costa Vasconcelos[3] | 1988 | Brazil | Insect | Limatus Sp | Seroprevalence | 0 | 1 | 0.000 | CF | IgM,NAb |
| PF da Costa Vasconcelos[3] | 1988 | Brazil | Insect | Mansonia Sp. | Seroprevalence | 0 | 1 | 0.000 | CF | IgM,NAb |
| PF da Costa Vasconcelos[3] | 1988 | Brazil | Insect | Psorophora cingulata | Seroprevalence | 0 | 1 | 0.000 | CF | IgM,NAb |
| PF da Costa Vasconcelos[3] | 1988 | Brazil | Insect | Psorophora ferox | Seroprevalence | 0 | 1 | 0.000 | CF | IgM,NAb |
| PF da Costa Vasconcelos[3] | 1988 | Brazil | Insect | Sabethes glaucodaemon | Seroprevalence | 0 | 1 | 0.000 | CF | IgM,NAb |
| PF da Costa Vasconcelos[3] | 1988 | Brazil | Insect | Uranotaenia Sp | Seroprevalence | 0 | 1 | 0.000 | CF | IgM,NAb |
| PF da Costa Vasconcelos[3] | 1988 | Brazil | Insect | Wyeomiya Sp. | Seroprevalence | 0 | 1 | 0.000 | CF | IgM,NAb |

| | | | | | | | | | | |
|---|---|---|---|---|---|---|---|---|---|---|
| Scott Medlin[4] | 2007 | Costa Rica | Non-human animal | Bradypus variegatus | Seroprevalence | 0 | 94 | 0.000 | HAI | NAb |
| Scott Medlin[4] | 2007 | Costa Rica | Non-human animal | Choloepus hoffmanni | Seroprevalence | 0 | 94 | 0.000 | HAI | NAb |
| Ana Cecilia Ribeiro Cruz[5] | 2007 | Brazil | Non-human animal | Conopophage roberti | Seroprevalence | 0 | 1 | 0.000 | HAI | NAb |
| Ana Cecilia Ribeiro Cruz[5] | 2007 | Brazil | Non-human animal | Didelphis marsupialis | Seroprevalence | 0 | 5 | 0.000 | HAI | NAb |
| Ana Cecilia Ribeiro Cruz[5] | 2007 | Brazil | Non-human animal | Geotrygon montana | Seroprevalence | 0 | 1 | 0.000 | HAI | NAb |
| Ana Cecilia Ribeiro Cruz[5] | 2007 | Brazil | Non-human animal | Myrmothera companisona | Seroprevalence | 0 | 1 | 0.000 | HAI | NAb |
| Ana Cecilia Ribeiro Cruz[5] | 2007 | Brazil | Non-human animal | Phlegopsis n. pareensis | Seroprevalence | 0 | 72 | 0.000 | HAI | NAb |
| Ana Cecilia Ribeiro Cruz[5] | 2007 | Brazil | Non-human animal | Proechimis guianensis | Seroprevalence | 0 | 2 | 0.000 | HAI | NAb |
| Ana Cecilia Ribeiro Cruz[5] | 2007 | Brazil | Non-human animal | Schiffornis tudinus | Seroprevalence | 0 | 1 | 0.000 | HAI | NAb |
| Ana Cecilia Ribeiro Cruz[5] | 2007 | Brazil | Non-human animal | Thamnophilus aethiops | Seroprevalence | 0 | 1 | 0.000 | HAI | NAb |
| Ana Cecilia Ribeiro Cruz[5] | 2007 | Brazil | Non-human animal | Xiphorhynchus ocellatus | Seroprevalence | 0 | 1 | 0.000 | HAI | NAb |
| Plautino O. Laroque[6] | 2010 | Brazil | Non-human animal | Cebus flavius | Seroprevalence | 0 | 31 | 0.000 | HAI | NAb |
| Plautino O. Laroque[6] | 2010 | Brazil | Non-human animal | Cebus libidinosus | Seroprevalence | 28 | 100 | 0.280 | HAI | NAb |
| Paulo Mira Batista[7] | 2010 | Brazil | Non-human animal | Multiple | Seroprevalence | 5 | 65 | 0.077 | HAI | NR |
| Alex Pauvolid-Correa[8] | 2011 | Brazil | Non-human animal | Caiman species | Seroprevalence | 0 | 66 | 0.000 | PRNT | NAb |
| Alex Pauvolid-Correa[8] | 2011 | Brazil | Non-human animal | Equine species | Seroprevalence | 0 | 375 | 0.000 | PRNT | NAb |
| Alex Pauvolid-Correa[8] | 2011 | Brazil | Non-human animal | Sheep species | Seroprevalence | 1 | 232 | 0.004 | PRNT | NAb |
| Michael J. Turell[9] | 2011 | Peru | Non-human animal | Aotus nancymaae | Seroprevalence | 0 | 20 | 0.000 | ELISA | IgG,IgM |

| | | | | | | | | | | |
|---|---|---|---|---|---|---|---|---|---|---|
| Belgath Fernandes Cardoso[10] | 2012 | Brazil | Insect | Culex quinquefasciatus | Viral Prevalence | 8 | 387 | 0.021 | RT-PCR | S segment |
| Paulo Mira Batista[11] | 2013 | Brazil | Non-human animal | Non-human primates | Seroprevalence | 3 | 48 | 0.062 | HAI | NAb |
| Paulo Mira Batista[11] | 2013 | Brazil | Non-human animal | Non-human primates | Seroprevalence | 3 | 48 | 0.062 | HAI | NAb |
| Paulo Mira Batista[11] | 2013 | Brazil | Non-human animal | Non-human primates | Seroprevalence | 3 | 48 | 0.062 | HAI | NAb |
| Paulo Mira Batista[11] | 2013 | Brazil | Non-human animal | Non-human primates | Seroprevalence | 3 | 48 | 0.062 | HAI | NAb |
| Paulo Mira Batista[11] | 2013 | Brazil | Non-human animal | Non-human primates | Seroprevalence | 3 | 48 | 0.062 | HAI | NAb |
| Paulo Mira Batista[11] | 2013 | Brazil | Non-human animal | Non-human primates | Seroprevalence | 3 | 48 | 0.062 | HAI | NAb |
| Paulo Mira Batista[11] | 2013 | Brazil | Non-human animal | Non-human primates | Seroprevalence | 3 | 48 | 0.062 | HAI | NAb |
| Paulo Mira Batista[11] | 2013 | Brazil | Non-human animal | Non-human primates | Seroprevalence | 3 | 48 | 0.062 | HAI | NAb |
| Paulo Mira Batista[11] | 2013 | Brazil | Non-human animal | Non-human primates | Seroprevalence | 3 | 48 | 0.062 | HAI | NAb |
| Jordam Pereira-Silva [12] | 2016 | Brazil | Insect | Multiple | Viral Prevalence | 3 | 671 | 0.004 | RT-PCR | S segment |
| Laura Tauro[13] | 2017 | Brazil | Insect | Aedes aegypti | Viral Prevalence | 0 | 26 | 0.000 | RT-PCR | NR |
| Laura Tauro[13] | 2017 | Brazil | Insect | Culex quinquefasciatus | Viral Prevalence | 0 | 99 | 0.000 | RT-PCR | NR |
| Raquel da Silva Ferreira[14] | 2018 | Brazil | Insect | Aedes aegypti | Viral Prevalence | 1 | 84 | 0.012 | RT-PCR | S segment |
| Raquel da Silva Ferreira[14] | 2018 | Brazil | Insect | Culex Spp. | Viral Prevalence | 0 | 3 | 0.000 | RT-PCR | S segment |
| Raquel da Silva Ferreira[14] | 2018 | Brazil | Insect | Culex quinquefasciatus | Viral Prevalence | 1 | 179 | 0.006 | RT-PCR | S segment |
| Raquel da Silva Ferreira[14] | 2018 | Brazil | Insect | Psorophora albigenu | Viral Prevalence | 0 | 1 | 0.000 | RT-PCR | S segment |
| Helver Gonçalves Dias[15] | 2018 | Brazil | Non-human animal | Bos indicus/taurus | Seroprevalence | 3 | 40 | 0.075 | PRNT | NAb |

| | | | | | | | | | | |
|---|---|---|---|---|---|---|---|---|---|---|
| Helver Gonçalves Dias[15] | 2018 | Brazil | Non-human animal | Canis lupus familiaris | Seroprevalence | 3 | 30 | 0.100 | PRNT | NAb |
| Helver Gonçalves Dias[15] | 2018 | Brazil | Non-human animal | Equus ferus caballus | Seroprevalence | 0 | 35 | 0.000 | PRNT | NAb |
| Helver Dias[16] | 2018 | Brazil | Insect | Aedes Spp. | Viral Prevalence | 0 | 62 | 0.000 | RT-PCR | S segment |
| Helver Dias[16] | 2018 | Brazil | Insect | Aedes Spp. | Viral Prevalence | 0 | 62 | 0.000 | RT-PCR | S segment |
| Helver Dias[16] | 2018 | Brazil | Insect | Aedes aegypti | Viral Prevalence | 0 | 280 | 0.000 | RT-PCR | S segment |
| Helver Dias[16] | 2018 | Brazil | Insect | Aedes aegypti | Viral Prevalence | 0 | 280 | 0.000 | RT-PCR | S segment |
| Helver Dias[16] | 2018 | Brazil | Insect | Aedes albopictus | Viral Prevalence | 0 | 94 | 0.000 | RT-PCR | S segment |
| Helver Dias[16] | 2018 | Brazil | Insect | Aedes albopictus | Viral Prevalence | 0 | 94 | 0.000 | RT-PCR | S segment |
| Helver Dias[16] | 2018 | Brazil | Insect | Aedes scapularis | Viral Prevalence | 0 | 101 | 0.000 | RT-PCR | S segment |
| Helver Dias[16] | 2018 | Brazil | Insect | Aedes scapularis | Viral Prevalence | 0 | 101 | 0.000 | RT-PCR | S segment |
| Helver Dias[16] | 2018 | Brazil | Non-human animal | Alouatta caraya | Viral Prevalence | 0 | 3 | 0.000 | RT-PCR | S segment |
| Helver Dias[16] | 2018 | Brazil | Non-human animal | Aotus lemurinus | Viral Prevalence | 0 | 2 | 0.000 | RT-PCR | S segment |
| Helver Dias[16] | 2018 | Brazil | Non-human animal | Ateles marginatus | Viral Prevalence | 0 | 2 | 0.000 | RT-PCR | S segment |
| Helver Dias[16] | 2018 | Brazil | Non-human animal | Bos indicus/taurus | Viral Prevalence | 0 | 176 | 0.000 | RT-PCR | S segment |
| Helver Dias[16] | 2018 | Brazil | Non-human animal | Bos indicus/taurus | Viral Prevalence | 0 | 176 | 0.000 | RT-PCR | S segment |
| Helver Dias[16] | 2018 | Brazil | Non-human animal | Callithrix jacchus | Viral Prevalence | 0 | 4 | 0.000 | RT-PCR | S segment |
| Helver Dias[16] | 2018 | Brazil | Non-human animal | Canis lupus familiaris | Viral Prevalence | 0 | 174 | 0.000 | RT-PCR | S segment |
| Helver Dias[16] | 2018 | Brazil | Non-human animal | Canis lupus familiaris | Viral Prevalence | 0 | 174 | 0.000 | RT-PCR | S segment |
| Helver Dias[16] | 2018 | Brazil | Insect | Culex Spp. | Viral Prevalence | 0 | 21,168 | 0.000 | RT-PCR | S segment |

| | | | | | | | | | | |
|---|---|---|---|---|---|---|---|---|---|---|
| Helver Dias[16] | 2018 | Brazil | Insect | Culex Spp. | Viral Prevalence | 0 | 21,168 | 0.000 | RT-PCR | S segment |
| Helver Dias[16] | 2018 | Brazil | Insect | Culex nigripalpus | Viral Prevalence | 0 | 175 | 0.000 | RT-PCR | S segment |
| Helver Dias[16] | 2018 | Brazil | Insect | Culex quinquefasciatus | Viral Prevalence | 0 | 75 | 0.000 | RT-PCR | S segment |
| Helver Dias[16] | 2018 | Brazil | Non-human animal | Didelphis albiventris | Viral Prevalence | 0 | 73 | 0.000 | RT-PCR | S segment |
| Helver Dias[16] | 2018 | Brazil | Non-human animal | Didelphis albiventris | Viral Prevalence | 0 | 73 | 0.000 | RT-PCR | S segment |
| Helver Dias[16] | 2018 | Brazil | Non-human animal | Didelphis aurita | Viral Prevalence | 0 | 3 | 0.000 | RT-PCR | S segment |
| Helver Dias[16] | 2018 | Brazil | Non-human animal | Equus ferus caballus | Viral Prevalence | 0 | 160 | 0.000 | RT-PCR | S segment |
| Helver Dias[16] | 2018 | Brazil | Non-human animal | Equus ferus caballus | Viral Prevalence | 0 | 160 | 0.000 | RT-PCR | S segment |
| Helver Dias[16] | 2018 | Brazil | Non-human animal | Felis silvestris catus | Viral Prevalence | 0 | 85 | 0.000 | RT-PCR | S segment |
| Helver Dias[16] | 2018 | Brazil | Non-human animal | Felis silvestris catus | Viral Prevalence | 0 | 85 | 0.000 | RT-PCR | S segment |
| Helver Dias[16] | 2018 | Brazil | Insect | Haemagogus Sp. | Viral Prevalence | 0 | 11 | 0.000 | RT-PCR | S segment |
| Helver Dias[16] | 2018 | Brazil | Insect | Haemagogus janthinomys | Viral Prevalence | 0 | 52 | 0.000 | RT-PCR | S segment |
| Helver Dias[16] | 2018 | Brazil | Insect | Haemagogus janthinomys | Viral Prevalence | 0 | 52 | 0.000 | RT-PCR | S segment |
| Helver Dias[16] | 2018 | Brazil | Insect | Haemagogus leucocelaenus | Viral Prevalence | 0 | 22 | 0.000 | RT-PCR | S segment |
| Helver Dias[16] | 2018 | Brazil | Insect | Haemagogus leucocelaenus | Viral Prevalence | 0 | 22 | 0.000 | RT-PCR | S segment |
| Helver Dias[16] | 2018 | Brazil | Insect | Mansonia Sp. | Viral Prevalence | 0 | 34 | 0.000 | RT-PCR | S segment |
| Helver Dias[16] | 2018 | Brazil | Insect | Mansonia Sp. | Viral Prevalence | 0 | 34 | 0.000 | RT-PCR | S segment |
| Helver Dias[16] | 2018 | Brazil | Non-human animal | Mico melanurus | Viral Prevalence | 0 | 29 | 0.000 | RT-PCR | S segment |

| | | | | | | | | | | |
|---|---|---|---|---|---|---|---|---|---|---|
| Helver Dias[16] | 2018 | Brazil | Insect | Multiple | Viral Prevalence | 0 | 22,931 | 0.000 | RT-PCR | S segment |
| Helver Dias[16] | 2018 | Brazil | Non-huma n animal | Nasua nasua | Viral Prevalence | 0 | 83 | 0.000 | RT-PCR | S segment |
| Helver Dias[16] | 2018 | Brazil | Non-huma n animal | Nasua nasua | Viral Prevalence | 0 | 83 | 0.000 | RT-PCR | S segment |
| Helver Dias[16] | 2018 | Brazil | Insect | Psorophora Spp. | Viral Prevalence | 0 | 94 | 0.000 | RT-PCR | S segment |
| Helver Dias[16] | 2018 | Brazil | Insect | Psorophora Spp. | Viral Prevalence | 0 | 94 | 0.000 | RT-PCR | S segment |
| Helver Dias[16] | 2018 | Brazil | Insect | Psorophora albigenu | Viral Prevalence | 0 | 134 | 0.000 | RT-PCR | S segment |
| Helver Dias[16] | 2018 | Brazil | Insect | Psorophora albigenu | Viral Prevalence | 0 | 134 | 0.000 | RT-PCR | S segment |
| Helver Dias[16] | 2018 | Brazil | Insect | Psorophora cilipes | Viral Prevalence | 0 | 38 | 0.000 | RT-PCR | S segment |
| Helver Dias[16] | 2018 | Brazil | Insect | Psorophora cilipes | Viral Prevalence | 0 | 38 | 0.000 | RT-PCR | S segment |
| Helver Dias[16] | 2018 | Brazil | Insect | Psorophora cingulata | Viral Prevalence | 0 | 39 | 0.000 | RT-PCR | S segment |
| Helver Dias[16] | 2018 | Brazil | Insect | Psorophora cingulata | Viral Prevalence | 0 | 39 | 0.000 | RT-PCR | S segment |
| Helver Dias[16] | 2018 | Brazil | Insect | Psorophora dimidiata | Viral Prevalence | 0 | 191 | 0.000 | RT-PCR | S segment |
| Helver Dias[16] | 2018 | Brazil | Insect | Psorophora dimidiata | Viral Prevalence | 0 | 191 | 0.000 | RT-PCR | S segment |
| Helver Dias[16] | 2018 | Brazil | Insect | Psorophora lanei | Viral Prevalence | 0 | 32 | 0.000 | RT-PCR | S segment |
| Helver Dias[16] | 2018 | Brazil | Insect | Psorophora lanei | Viral Prevalence | 0 | 32 | 0.000 | RT-PCR | S segment |
| Helver Dias[16] | 2018 | Brazil | Insect | Sabethes Spp. | Viral Prevalence | 0 | 16 | 0.000 | RT-PCR | S segment |
| Helver Dias[16] | 2018 | Brazil | Insect | Sabethes Spp. | Viral Prevalence | 0 | 16 | 0.000 | RT-PCR | S segment |
| Helver Dias[16] | 2018 | Brazil | Non-huma n animal | Sapajus apella | Viral Prevalence | 0 | 5 | 0.000 | RT-PCR | S segment |
| Helver Dias[16] | 2018 | Brazil | Non-huma n animal | Sapajus cay | Viral Prevalence | 0 | 11 | 0.000 | RT-PCR | S segment |

| | | | | | | | | | | |
|---|---|---|---|---|---|---|---|---|---|---|
| Helver Dias[16] | 2018 | Brazil | Insect | Wyeomiya Sp. | Viral Prevalence | 0 | 313 | 0.000 | RT-PCR | S segment |
| Helver Dias[16] | 2018 | Brazil | Insect | Wyeomiya Sp. | Viral Prevalence | 0 | 313 | 0.000 | RT-PCR | S segment |
| Luiz Henrique Maciel Feitoza[17] | 2020 | Brazil | Insect | Culicoides paraensis | Viral Prevalence | 0 | 271 | 0.000 | RT-PCR | S segment |
| Luiz Henrique Maciel Feitoza[17] | 2020 | Brazil | Insect | Culicoides paraensis | Viral Prevalence | 0 | 271 | 0.000 | RT-PCR | S segment |
| Luiz Henrique Maciel Feitoza[17] | 2020 | Brazil | Insect | Culicoides paraensis | Viral Prevalence | 0 | 271 | 0.000 | RT-PCR | S segment |
| Diego Michel Fernandes da Silva[18] | 2022 | Brazil | Insect | Aedes aegypti | Viral Prevalence | 0 | 1,570 | 0.000 | RT-PCR | NR |

#### Table C. Risk of bias breakdown for all studies.

| Author | Grouping Variable | Study Population (ORO | Study Species (ORO | Estimate Type | Item 1 (JBI-M) | Item 5 (JBI-M) | Item 2 (JBI-A) | Item 3 (JBI-A) | Item 4 (JBI-A) | Item 6 (JBI-A) | Item 7 (JBI-A) | Item 8B (JBI-A) | Item 8A (JBI-A) | JBI-A Outputs |
|---|---|---|---|---|---|---|---|---|---|---|---|---|---|---|

| | | | | | | | | | | | | | | |
|---|---|---|---|---|---|---|---|---|---|---|---|---|---|---|
| | | **V only)** | **V only)** | | | | | | | | | | | |
| Pedro P. Alvarez | Overall | Human | Homo sapiens | Seroprevalence | Yes | Unclear | No | Yes | Yes | Yes | Yes | No | Yes | Moderate |
| Juana del Valle-Mendoza | Overall | Human | Homo sapiens | Viral Prevalence | No | Unclear | No | No | No | Yes | Yes | No | Yes | High |
| F. P. Pinheiro | Species | Human | Homo sapiens | Seroprevalence | Yes | Unclear | No | Yes | Yes | Yes | Yes | No | Yes | Moderate |
| Brett Forshey | Geography | Human | Homo sapiens | Viral Prevalence | Yes | Yes | Yes | Yes | Yes | Yes | Yes | Yes | Yes | Low |
| Brett Forshey | Geography | Human | Homo sapiens | Viral Prevalence | Yes | Yes | Yes | Yes | Yes | Yes | Yes | Yes | Yes | Low |
| Brett Forshey | Geography | Human | Homo sapiens | Viral Prevalence | Yes | Yes | Yes | Yes | Yes | Yes | Yes | Yes | Yes | Low |
| Brett Forshey | Geography | Human | Homo sapiens | Viral Prevalence | Yes | Yes | Yes | Yes | Yes | Yes | Yes | Yes | Yes | Low |
| Brett Forshey | Geography | Human | Homo sapiens | Viral Prevalence | Yes | Yes | Yes | Yes | Yes | Yes | Yes | Yes | Yes | Low |
| Brett Forshey | Geography | Human | Homo sapiens | Viral Prevalence | Yes | Yes | Yes | Yes | Yes | Yes | Yes | Yes | Yes | Low |
| Brett Forshey | Geography | Human | Homo sapiens | Viral Prevalence | Yes | Yes | Yes | Yes | Yes | Yes | Yes | Yes | Yes | Low |
| Brett Forshey | Geography | Human | Homo sapiens | Viral Prevalence | Yes | Yes | Yes | Yes | Yes | Yes | Yes | Yes | Yes | Low |
| Brett Forshey | Geography | Human | Homo sapiens | Viral Prevalence | Yes | Yes | Yes | Yes | Yes | Yes | Yes | Yes | Yes | Low |
| Brett Forshey | Geography | Human | Homo sapiens | Viral Prevalence | Yes | Yes | Yes | Yes | Yes | Yes | Yes | Yes | Yes | Low |
| Brett Forshey | Geography | Human | Homo sapiens | Viral Prevalence | Yes | Yes | Yes | Yes | Yes | Yes | Yes | Yes | Yes | Low |
| Brett Forshey | Geography | Human | Homo sapiens | Viral Prevalence | Yes | Yes | Yes | Yes | Yes | Yes | Yes | Yes | Yes | Low |
| Brett Forshey | Geography | Human | Homo sapiens | Viral Prevalence | Yes | Yes | Yes | Yes | Yes | Yes | Yes | Yes | Yes | Low |

| | | | | | | | | | | | | | | |
|---|---|---|---|---|---|---|---|---|---|---|---|---|---|---|
| Brett Forshey | Gender | Human | Homo sapiens | Viral Prevalence | Yes | Yes | Yes | Yes | Yes | Yes | Yes | Yes | Yes | Low |
| Brett Forshey | Gender | Human | Homo sapiens | Viral Prevalence | Yes | Yes | Yes | Yes | Yes | Yes | Yes | Yes | Yes | Low |
| Juliana Gil-Mora | Geography | Human | Homo sapiens | Seroprevalence | Yes | No | Yes | Yes | Yes | Yes | Yes | Yes | Yes | Low |
| Juliana Gil-Mora | Geography | Human | Homo sapiens | Seroprevalence | Yes | No | Yes | Yes | Yes | Yes | Yes | Yes | Yes | Low |
| Juliana Gil-Mora | Geography | Human | Homo sapiens | Seroprevalence | Yes | No | Yes | No | Yes | Yes | Yes | Yes | Yes | Moderate |
| Juliana Gil-Mora | Geography | Human | Homo sapiens | Seroprevalence | Yes | No | Yes | No | Yes | Yes | Yes | Yes | Yes | Moderate |
| M.C. de Souza Costa | Gender | Human | Homo sapiens | Viral Prevalence | Yes | No | Yes | Yes | Yes | Yes | Yes | Yes | Yes | Low |
| M.C. de Souza Costa | Gender | Human | Homo sapiens | Viral Prevalence | Yes | No | Yes | Yes | Yes | Yes | Yes | Yes | Yes | Low |
| Vanessa L. Carvalho | Overall | Human | Homo sapiens | Seroprevalence | No | Unclear | Yes | No | No | Yes | Yes | No | Yes | High |
| Sara Castro | Test type | Human | Homo sapiens | Viral Prevalence | Yes | No | Yes | No | Yes | Yes | Yes | Yes | Yes | Moderate |
| Sara Castro | Test type | Human | Homo sapiens | Seroprevalence | Yes | No | Yes | No | Yes | Yes | Yes | Yes | Yes | Moderate |
| Valquiria do Carmo Alves Martins | Overall | Human | Homo sapiens | Viral Prevalence | No | Yes | Yes | Yes | Yes | Yes | Yes | No | Yes | Moderate |
| Maria Garcia | Overall | Human | Homo sapiens | Seroprevalence | Yes | Yes | Yes | Yes | No | Yes | Yes | Yes | Yes | Low |
| Maria Garcia | Overall | Human | Homo sapiens | Viral Prevalence | Yes | Yes | Yes | Yes | No | Yes | Yes | Yes | Yes | Low |
| Maria Garcia | Overall | Human | Homo sapiens | Viral Prevalence | Yes | Yes | Yes | Yes | No | Yes | Yes | Yes | Yes | Low |

| | | | | | | | | | | | | | | |
|---|---|---|---|---|---|---|---|---|---|---|---|---|---|---|
| Michele S. Bastos | Overall | Human | Homo sapiens | Viral Prevalence | Yes | Yes | Yes | Yes | Yes | Yes | Yes | Yes | Yes | Low |
| Belgath Fernandes Cardoso | Gender | Human | Homo sapiens | Viral Prevalence | No | Yes | No | Yes | Yes | Yes | Yes | No | Yes | High |
| Belgath Fernandes Cardoso | Gender | Human | Homo sapiens | Viral Prevalence | No | Yes | No | Yes | Yes | Yes | Yes | No | Yes | High |
| Regina Maria Pinto De Figueiredo | Overall | Human | Homo sapiens | Seroprevalence | No | Yes | Yes | No | Yes | Yes | Yes | No | Yes | High |
| Marcio Nunes | Timeframe | Human | Homo sapiens | Seroprevalence | No | Unclear | No | Yes | No | Yes | Yes | No | Yes | High |
| Marcio Nunes | Timeframe | Human | Homo sapiens | Seroprevalence | No | Unclear | No | Yes | No | Yes | Yes | No | Yes | High |
| Marcio Nunes | Timeframe | Human | Homo sapiens | Seroprevalence | No | Unclear | No | Yes | No | Yes | Yes | No | Yes | High |
| Marcio Nunes | Timeframe | Human | Homo sapiens | Seroprevalence | No | Unclear | No | Yes | No | Yes | Yes | No | Yes | High |
| Marcio Nunes | Timeframe | Human | Homo sapiens | Seroprevalence | No | Unclear | No | Yes | No | Yes | Yes | No | Yes | High |
| Carlos Alva-Urcia | Gender | Human | Homo sapiens | Viral Prevalence | Yes | No | Yes | No | Yes | Yes | Yes | Yes | Yes | Moderate |
| Carlos Alva-Urcia | Gender | Human | Homo sapiens | Viral Prevalence | Yes | No | Yes | No | Yes | Yes | Yes | Yes | Yes | Moderate |
| Amélia Rosa | Gender | Human | Homo sapiens | Seroprevalence | Yes | Yes | Yes | Yes | Yes | Yes | Yes | Yes | Yes | Low |
| Amélia Rosa | Gender | Human | Homo sapiens | Seroprevalence | Yes | Yes | Yes | Yes | Yes | Yes | Yes | Yes | Yes | Low |
| Ronaldo B. Freitas | Gender | Human | Homo sapiens | Seroprevalence | Yes | Unclear | Yes | Yes | Yes | Yes | Yes | Yes | Yes | Low |

| | | | | | | | | | | | | | | |
|---|---|---|---|---|---|---|---|---|---|---|---|---|---|---|
| Ronaldo B. Freitas | Gender | Human | Homo sapiens | Seroprevalence | Yes | Unclear | Yes | Yes | Yes | Yes | Yes | Yes | Yes | Low |
| Ronaldo B. Freitas | Gender | Human | Homo sapiens | Seroprevalence | Yes | Unclear | Yes | Yes | Yes | Yes | Yes | Yes | Yes | Low |
| Ronaldo B. Freitas | Gender | Human | Homo sapiens | Seroprevalence | Yes | Unclear | Yes | Yes | Yes | Yes | Yes | Yes | Yes | Low |
| Ronaldo B. Freitas | Gender | Human | Homo sapiens | Seroprevalence | Yes | Unclear | Yes | Yes | Yes | Yes | Yes | Yes | Yes | Low |
| Ronaldo B. Freitas | Gender | Human | Homo sapiens | Seroprevalence | Yes | Unclear | Yes | Yes | Yes | Yes | Yes | Yes | Yes | Low |
| Ronaldo B. Freitas | Gender | Human | Homo sapiens | Seroprevalence | Yes | Unclear | Yes | No | Yes | Yes | Yes | Yes | Yes | Moderate |
| Ronaldo B. Freitas | Gender | Human | Homo sapiens | Seroprevalence | Yes | Unclear | Yes | No | Yes | Yes | Yes | Yes | Yes | Moderate |
| Ronaldo B. Freitas | Gender | Human | Homo sapiens | Seroprevalence | Yes | Unclear | Yes | No | Yes | Yes | Yes | Yes | Yes | Moderate |
| Ronaldo B. Freitas | Gender | Human | Homo sapiens | Seroprevalence | Yes | Unclear | Yes | No | Yes | Yes | Yes | Yes | Yes | Moderate |
| Ronaldo B. Freitas | Gender | Human | Homo sapiens | Seroprevalence | Yes | Unclear | Yes | Yes | Yes | Yes | Yes | Yes | Yes | Low |
| Ronaldo B. Freitas | Gender | Human | Homo sapiens | Seroprevalence | Yes | Unclear | Yes | Yes | Yes | Yes | Yes | Yes | Yes | Low |
| Ronaldo B. Freitas | Gender | Human | Homo sapiens | Seroprevalence | Yes | Unclear | Yes | No | Yes | Yes | Yes | Yes | Yes | Moderate |
| Ronaldo B. Freitas | Gender | Human | Homo sapiens | Seroprevalence | Yes | Unclear | Yes | No | Yes | Yes | Yes | Yes | Yes | Moderate |
| Ronaldo B. Freitas | Test type | Human | Homo sapiens | Seroprevalence | Yes | Unclear | Yes | Yes | Yes | Yes | Yes | Yes | Yes | Low |
| Ronaldo B. Freitas | Test type | Human | Homo sapiens | Viral Prevalence | Yes | Unclear | Yes | Yes | Yes | Yes | Yes | Yes | Yes | Low |
| Ronaldo B. Freitas | Test type | Human | Homo sapiens | Seroprevalence | Yes | Unclear | Yes | No | Yes | Yes | Yes | Yes | Yes | Moderate |
| Ronaldo B. Freitas | Test type | Human | Homo sapiens | Viral Prevalence | Yes | Unclear | Yes | No | Yes | Yes | Yes | Yes | Yes | Moderate |

| | | | | | | | | | | | | | | |
|---|---|---|---|---|---|---|---|---|---|---|---|---|---|---|
| Ronaldo B. Freitas | Test type | Human | Homo sapiens | Seroprevalence | Yes | Unclear | Yes | No | Yes | Yes | Yes | Yes | Yes | Moderate |
| Ronaldo B. Freitas | Test type | Human | Homo sapiens | Viral Prevalence | Yes | Unclear | Yes | No | Yes | Yes | Yes | Yes | Yes | Moderate |
| Ronaldo B. Freitas | Test type | Human | Homo sapiens | Seroprevalence | Yes | Unclear | Yes | No | Yes | Yes | Yes | Yes | Yes | Moderate |
| Ronaldo B. Freitas | Test type | Human | Homo sapiens | Viral Prevalence | Yes | Unclear | Yes | No | Yes | Yes | Yes | Yes | Yes | Moderate |
| Ronaldo B. Freitas | Test type | Human | Homo sapiens | Seroprevalence | Yes | Unclear | Yes | No | Yes | Yes | Yes | Yes | Yes | Moderate |
| Ronaldo B. Freitas | Test type | Human | Homo sapiens | Viral Prevalence | Yes | Unclear | Yes | No | Yes | Yes | Yes | Yes | Yes | Moderate |
| Ronaldo B. Freitas | Test type | Human | Homo sapiens | Seroprevalence | Yes | Unclear | Yes | Yes | Yes | Yes | Yes | Yes | Yes | Low |
| Ronaldo B. Freitas | Test type | Human | Homo sapiens | Viral Prevalence | Yes | Unclear | Yes | Yes | Yes | Yes | Yes | Yes | Yes | Low |
| Ronaldo B. Freitas | Test type | Human | Homo sapiens | Seroprevalence | Yes | Unclear | Yes | No | Yes | Yes | Yes | Yes | Yes | Moderate |
| Ronaldo B. Freitas | Test type | Human | Homo sapiens | Viral Prevalence | Yes | Unclear | Yes | No | Yes | Yes | Yes | Yes | Yes | Moderate |
| Ronaldo B. Freitas | Test type | Human | Homo sapiens | Seroprevalence | Yes | Unclear | Yes | No | Yes | Yes | Yes | Yes | Yes | Moderate |
| Ronaldo B. Freitas | Test type | Human | Homo sapiens | Viral Prevalence | Yes | Unclear | Yes | No | Yes | Yes | Yes | Yes | Yes | Moderate |
| Ronaldo B. Freitas | Test type | Human | Homo sapiens | Seroprevalence | Yes | Unclear | Yes | No | Yes | Yes | Yes | Yes | Yes | Moderate |
| Ronaldo B. Freitas | Test type | Human | Homo sapiens | Viral Prevalence | Yes | Unclear | Yes | No | Yes | Yes | Yes | Yes | Yes | Moderate |
| James LeDuc | Timeframe | Human | Homo sapiens | Seroprevalence | No | Yes | No | No | Yes | Yes | Yes | No | Yes | High |
| James LeDuc | Timeframe | Human | Homo sapiens | Seroprevalence | No | Yes | No | Yes | Yes | Yes | Yes | No | Yes | High |
| James LeDuc | Timeframe | Human | Homo sapiens | Seroprevalence | No | Yes | No | Yes | Yes | Yes | Yes | No | Yes | High |

| | | | | | | | | | | | | | | |
|---|---|---|---|---|---|---|---|---|---|---|---|---|---|---|
| Kathy Baisley | Gender | Human | Homo sapiens | Seroprevalence | Yes | Yes | Yes | Yes | Yes | Yes | Yes | Yes | Yes | Low |
| Kathy Baisley | Gender | Human | Homo sapiens | Seroprevalence | Yes | Yes | Yes | Yes | Yes | Yes | Yes | Yes | Yes | Low |
| Carlos Silva-Ramos | Overall | Human | Homo sapiens | Viral Prevalence | Yes | No | Yes | No | Yes | Yes | Yes | Yes | Yes | Moderate |
| Carlos Silva-Ramos | Overall | Human | Homo sapiens | Viral Prevalence | Yes | No | Yes | No | Yes | Yes | Yes | Yes | Yes | Moderate |
| Douglas M. Watts | Overall | Human | Homo sapiens | Seroprevalence | Yes | Yes | Yes | Yes | Yes | Yes | Yes | Yes | Yes | Low |
| Douglas M. Watts | Gender | Human | Homo sapiens | Seroprevalence | Yes | Yes | Yes | Yes | Yes | Yes | Yes | Yes | Yes | Low |
| Douglas M. Watts | Gender | Human | Homo sapiens | Seroprevalence | Yes | Yes | Yes | Yes | Yes | Yes | Yes | Yes | Yes | Low |
| Stephen Manock | Test type | Human | Homo sapiens | Seroprevalence | No | Yes | Yes | Yes | Yes | Yes | Yes | No | Yes | Moderate |
| Stephen Manock | Test type | Human | Homo sapiens | Seroprevalence | No | Yes | Yes | Yes | Yes | Yes | Yes | No | Yes | Moderate |
| Wilmer Silva-Caso | Gender | Human | Homo sapiens | Viral Prevalence | No | Yes | No | Yes | Yes | Yes | Yes | No | Yes | High |
| Wilmer Silva-Caso | Gender | Human | Homo sapiens | Viral Prevalence | No | Yes | No | Yes | Yes | Yes | Yes | No | Yes | High |
| Felipe Naveca | Overall | Human | Homo sapiens | Viral Prevalence | Yes | Unclear | Yes | No | No | Yes | Yes | Yes | Yes | Moderate |
| Cassiano Junior Saatkamp | Overall | Human | Homo sapiens | Viral Prevalence | Yes | Unclear | Yes | No | Yes | Yes | Yes | Yes | Yes | Moderate |
| Janeth Aracely Ramirez Pavon | Overall | Human | Homo sapiens | Viral Prevalence | Yes | No | Yes | No | Yes | Yes | Yes | Yes | Yes | Moderate |

| | | | | | | | | | | | | | | |
|---|---|---|---|---|---|---|---|---|---|---|---|---|---|---|
| Helena Vasconcelos | Test type | Human | Homo sapiens | Seroprevalence | No | Unclear | Yes | Yes | Yes | Yes | Yes | No | Yes | Moderate |
| Helena Vasconcelos | Test type | Human | Homo sapiens | Seroprevalence | No | Unclear | Yes | Yes | Yes | Yes | Yes | No | Yes | Moderate |
| Helena Vasconcelos | Geography | Human | Homo sapiens | Seroprevalence | No | Unclear | Yes | Yes | Yes | Yes | Yes | No | Yes | Moderate |
| Helena Vasconcelos | Geography | Human | Homo sapiens | Seroprevalence | No | Unclear | Yes | No | Yes | Yes | Yes | No | Yes | High |
| Maria Paula Mourão | Overall | Human | Homo sapiens | Seroprevalence | Yes | No | Yes | Yes | Yes | Yes | Yes | Yes | Yes | Low |
| Johanna Martins-Luna | Overall | Human | Homo sapiens | Viral Prevalence | Yes | Yes | Yes | Yes | Yes | Yes | Yes | Yes | Yes | Low |
| Johanna Martins-Luna | Gender | Human | Homo sapiens | Viral Prevalence | Yes | Yes | Yes | Yes | Yes | Yes | Yes | Yes | Yes | Low |
| Johanna Martins-Luna | Gender | Human | Homo sapiens | Viral Prevalence | Yes | Yes | Yes | Yes | Yes | Yes | Yes | Yes | Yes | Low |
| Karl Ciuoderis | Test type | Human | Homo sapiens | Viral Prevalence | Yes | No | Yes | Yes | Yes | Yes | Yes | Yes | Yes | Low |
| Karl Ciuoderis | Test type | Human | Homo sapiens | Seroprevalence | Yes | No | Yes | Yes | Yes | Yes | Yes | Yes | Yes | Low |
| Karl Ciuoderis | Test type | Human | Homo sapiens | Seroprevalence | Yes | No | Yes | Yes | Yes | Yes | Yes | Yes | Yes | Low |
| Karl Ciuoderis | Test type | Human | Homo sapiens | Viral Prevalence | Yes | No | Yes | No | Yes | Yes | Yes | Yes | Yes | Moderate |
| Karl Ciuoderis | Test type | Human | Homo sapiens | Seroprevalence | Yes | No | Yes | No | Yes | Yes | Yes | Yes | Yes | Moderate |
| Karl Ciuoderis | Test type | Human | Homo sapiens | Viral Prevalence | Yes | No | Yes | No | Yes | Yes | Yes | Yes | Yes | Moderate |
| Karl Ciuoderis | Test type | Human | Homo sapiens | Seroprevalence | Yes | No | Yes | No | Yes | Yes | Yes | Yes | Yes | Moderate |
| Karl Ciuoderis | Test type | Human | Homo sapiens | Viral Prevalence | Yes | No | Yes | Yes | Yes | Yes | Yes | Yes | Yes | Low |

| | | | | | | | | | | | | | | |
|---|---|---|---|---|---|---|---|---|---|---|---|---|---|---|
| Karl Ciuoderis | Test type | Human | Homo sapiens | Seroprevalence | Yes | No | Yes | Yes | Yes | Yes | Yes | Yes | Yes | Low |
| Karl Ciuoderis | Test type | Human | Homo sapiens | Viral Prevalence | Yes | No | Yes | Yes | Yes | Yes | Yes | Yes | Yes | Low |
| Karl Ciuoderis | Test type | Human | Homo sapiens | Seroprevalence | Yes | No | Yes | Yes | Yes | Yes | Yes | Yes | Yes | Low |
| Emma L. Wise | Overall | Human | Homo sapiens | Viral Prevalence | No | Unclear | Yes | Yes | Yes | Yes | Yes | No | Yes | Moderate |
| Emma L. Wise | Overall | Human | Homo sapiens | Viral Prevalence | No | Unclear | Yes | No | Yes | Yes | Yes | No | Yes | High |
| Larissa Moraes dos Santos Fonseca | Overall | Human | Homo sapiens | Viral Prevalence | Yes | Unclear | Yes | No | No | Yes | Yes | Yes | Yes | Moderate |
| Valdinete Alves do Nascimento | Overall | Human | Homo sapiens | Viral Prevalence | Yes | Unclear | Yes | Yes | No | Yes | Yes | Yes | Yes | Low |
| Raquel Curtinhas de Lima | Overall | Human | Homo sapiens | Seroprevalence | Yes | Yes | Yes | Yes | Yes | Yes | Yes | Yes | Yes | Low |
| Raquel Curtinhas de Lima | Overall | Human | Homo sapiens | Viral Prevalence | Yes | Yes | Yes | Yes | Yes | Yes | Yes | Yes | Yes | Low |
| Hilda Durango-Chavez | Overall | Human | Homo sapiens | Viral Prevalence | Yes | Yes | Yes | Yes | Yes | Yes | Yes | Yes | Yes | Low |
| Wilmer Silva-Caso | Overall | Human | Homo sapiens | Viral Prevalence | Yes | Unclear | Yes | Yes | No | Yes | Yes | Yes | Yes | Low |
| Johanna Martins-Luna | Overall | Human | Homo sapiens | Viral Prevalence | Yes | Unclear | Yes | Yes | No | Yes | Yes | Yes | Yes | Low |
| Maha Elbadry | Overall | Human | Homo sapiens | Viral Prevalence | Yes | Unclear | Yes | Yes | Yes | Yes | Yes | Yes | Yes | Low |
| Mélanie Gaillet | Overall | Human | Homo sapiens | Viral Prevalence | No | Yes | Yes | No | Yes | Yes | Yes | No | Yes | High |

| | | | | | | | | | | | | | | |
|---|---|---|---|---|---|---|---|---|---|---|---|---|---|---|
| Hillquias Monteiro Moreira | Overall | Human | Homo sapiens | Viral Prevalence | Yes | Unclear | Yes | Yes | No | Yes | Yes | Yes | Yes | Low |
| José Tavares-Neto | Overall | Human | Homo sapiens | Seroprevalence | No | Unclear | No | Yes | Yes | Yes | Yes | No | Yes | High |
| José Tavares-Neto | Overall | Human | Homo sapiens | Seroprevalence | No | Unclear | Yes | Yes | Yes | Yes | Yes | No | Yes | Moderate |
| Jackson Alves da Silva Queiroz | Overall | Human | Homo sapiens | Viral Prevalence | Yes | No | Yes | Yes | No | Yes | Yes | Yes | Yes | Low |
| Barbara Batista Salgado | Overall | Human | Homo sapiens | Seroprevalence | No | No | Yes | Yes | Yes | Yes | Yes | No | Yes | Moderate |
| Luiz Henrique Gonçalves Maciel | Overall | Human | Homo sapiens | Viral Prevalence | Yes | Yes | Yes | Yes | Yes | Yes | Yes | Yes | Yes | Low |
| Carlos Borborema | Geography | Human | Homo sapiens | Seroprevalence | Yes | Unclear | Yes | No | No | Yes | Yes | Yes | Yes | Moderate |
| Carlos Borborema | Geography | Human | Homo sapiens | Seroprevalence | Yes | Unclear | Yes | No | No | Yes | Yes | Yes | Yes | Moderate |
| Carlos Borborema | Geography | Human | Homo sapiens | Seroprevalence | Yes | Unclear | Yes | No | No | Yes | Yes | Yes | Yes | Moderate |
| Carlos Borborema | Geography | Human | Homo sapiens | Seroprevalence | Yes | Unclear | Yes | Yes | No | Yes | Yes | Yes | Yes | Low |
| Carlos Borborema | Geography | Human | Homo sapiens | Seroprevalence | Yes | Unclear | Yes | No | No | Yes | Yes | Yes | Yes | Moderate |
| Carlos Borborema | Geography | Human | Homo sapiens | Seroprevalence | Yes | Unclear | Yes | No | No | Yes | Yes | Yes | Yes | Moderate |
| Carlos Borborema | Geography | Human | Homo sapiens | Seroprevalence | Yes | Unclear | Yes | No | No | Yes | Yes | Yes | Yes | Moderate |

| | | | | | | | | | | | | | | |
|---|---|---|---|---|---|---|---|---|---|---|---|---|---|---|
| Carlos Borborema | Geography | Human | Homo sapiens | Seroprevalence | Yes | Unclear | Yes | No | No | Yes | Yes | Yes | Yes | Moderate |
| Carlos Borborema | Geography | Human | Homo sapiens | Seroprevalence | Yes | Unclear | Yes | No | No | Yes | Yes | Yes | Yes | Moderate |
| Carlos Borborema | Geography | Human | Homo sapiens | Seroprevalence | Yes | Unclear | Yes | Yes | No | Yes | Yes | Yes | Yes | Low |
| Carlos Borborema | Geography | Human | Homo sapiens | Seroprevalence | Yes | Unclear | Yes | No | No | Yes | Yes | Yes | Yes | Moderate |
| Carlos Borborema | Geography | Human | Homo sapiens | Seroprevalence | Yes | Unclear | Yes | No | No | Yes | Yes | Yes | Yes | Moderate |
| Carlos Borborema | Geography | Human | Homo sapiens | Seroprevalence | Yes | Unclear | No | No | No | Yes | Yes | No | Yes | High |
| Carlos Borborema | Geography | Human | Homo sapiens | Seroprevalence | Yes | Unclear | No | No | No | Yes | Yes | No | Yes | High |
| Pedro Fernando da Costa Vasconcelos | Gender | Human | Homo sapiens | Seroprevalence | Unclear | Yes | Yes | Yes | Yes | Yes | Yes | No | Yes | High |
| Pedro Fernando da Costa Vasconcelos | Gender | Human | Homo sapiens | Seroprevalence | Unclear | Yes | Yes | No | Yes | Yes | Yes | No | Yes | High |
| Diego Michel Fernandes da Silva | Overall | Human | Homo sapiens | Viral Prevalence | Yes | Unclear | Yes | No | Yes | Yes | Yes | Yes | Yes | Moderate |
| Marco Coaguila | Overall | Human | Homo sapiens | Seroprevalence | Yes | Unclear | Yes | Yes | No | Yes | Yes | Yes | Yes | Low |
| Raimunda do Socorro da Silva Azevedo | Gender | Human | Homo sapiens | Seroprevalence | Yes | Yes | No | No | Yes | Yes | Yes | No | Yes | High |
| Raimunda do Socorro da Silva | Gender | Human | Homo sapiens | Seroprevalence | Yes | Yes | No | No | Yes | Yes | Yes | No | Yes | High |

| | | | | | | | | | | | | | | |
|---|---|---|---|---|---|---|---|---|---|---|---|---|---|---|
| Azevedo | | | | | | | | | | | | | | |
| Raimunda do Socorro da Silva Azevedo | Overall | Human | Homo sapiens | Seroprevalence | Yes | Yes | No | Yes | Yes | Yes | Yes | No | Yes | Moderate |
| Gabriel Scachetti | Overall | Human | Homo sapiens | Viral Prevalence | No | Unclear | No | No | No | Yes | Yes | No | Yes | High |
| Helver Dias | Species | Human | Homo sapiens | Viral Prevalence | Yes | No | Yes | Yes | Yes | Yes | Yes | Yes | Yes | Low |
| Tung Gia Phan | Overall | Human | Homo sapiens | Viral Prevalence | Unclear | Unclear | No | No | No | Yes | Yes | No | Yes | High |
| Vivaldo Gomes da Costa | Test type | Human | Homo sapiens | Seroprevalence | Unclear | Yes | No | Yes | Yes | Yes | Yes | No | Yes | High |
| Vivaldo Gomes da Costa | Test type | Human | Homo sapiens | Seroprevalence | Unclear | Yes | No | Yes | Yes | Yes | Yes | No | Yes | High |
| Ana Carolina Bernardes Terzian | Overall | Human | Homo sapiens | Viral Prevalence | Yes | Unclear | Yes | No | Yes | Yes | Yes | Yes | Yes | Moderate |
| Liliana Sanchez-Lerma | Overall | Human | Homo sapiens | Viral Prevalence | No | No | Yes | Yes | Yes | Yes | Yes | No | Yes | Moderate |
| Juana del Valle-Mendoza | Gender | Human | Homo sapiens | Viral Prevalence | Yes | Yes | Yes | No | Yes | Yes | Yes | Yes | Yes | Moderate |
| Juana del Valle-Mendoza | Gender | Human | Homo sapiens | Viral Prevalence | Yes | Yes | Yes | No | Yes | Yes | Yes | Yes | Yes | Moderate |
| Douglas Watts | Test type | Human | Homo sapiens | Seroprevalence | Yes | Unclear | No | No | No | Yes | Yes | No | Yes | High |

| | | | | | | | | | | | | | | |
|---|---|---|---|---|---|---|---|---|---|---|---|---|---|---|
| Douglas Watts | Test type | Human | Homo sapiens | Seroprevalence | Yes | Unclear | No | No | No | Yes | Yes | No | Yes | High |
| Ana Cecilia Ribeiro Cruz | Test type | Human | Homo sapiens | Seroprevalence | Yes | Unclear | Yes | Yes | No | Yes | Yes | Yes | Yes | Low |
| Ana Cecilia Ribeiro Cruz | Test type | Human | Homo sapiens | Seroprevalence | Yes | Unclear | Yes | Yes | No | Yes | Yes | Yes | Yes | Low |
| Laura Tauro | Overall | Insect | Culex quinquefasciatus | Viral Prevalence | Yes | Yes | Yes | No | No | Yes | Yes | Yes | Yes | Moderate |
| Laura Tauro | Overall | Insect | Aedes aegypti | Viral Prevalence | Yes | Yes | Yes | No | No | Yes | Yes | Yes | Yes | Moderate |
| F. P. Pinheiro | Species | Insect | Culicoides paraensis | Viral Prevalence | Yes | Unclear | No | Yes | No | Yes | Yes | No | Yes | Moderate |
| Belgath Fernandes Cardoso | Species | Insect | Culex quinquefasciatus | Viral Prevalence | Yes | Unclear | Yes | Yes | No | Yes | Yes | Yes | Yes | Low |
| Jordam Pereira-Silva | Overall | Insect | Multiple | Viral Prevalence | Yes | No | Yes | Yes | Yes | Yes | Yes | Yes | Yes | Low |
| Luiz Henrique Maciel Feitoza | Geography | Insect | Culicoides paraensis | Viral Prevalence | Yes | Unclear | No | No | Yes | Yes | Yes | No | Yes | High |
| Luiz Henrique Maciel Feitoza | Geography | Insect | Culicoides paraensis | Viral Prevalence | Yes | Unclear | No | No | Yes | Yes | Yes | No | Yes | High |
| Luiz Henrique Maciel Feitoza | Geography | Insect | Culicoides paraensis | Viral Prevalence | Yes | Unclear | No | Yes | Yes | Yes | Yes | No | Yes | Moderate |
| Raquel da Silva Ferreira | Species | Insect | Culex quinquefasciatus | Viral Prevalence | Yes | No | Yes | Yes | No | Yes | Yes | Yes | Yes | Low |

| | | | | | | | | | | | | | | |
|---|---|---|---|---|---|---|---|---|---|---|---|---|---|---|
| Raquel da Silva Ferreira | Species | Insect | Aedes aegypti | Viral Prevalence | Yes | No | Yes | No | No | Yes | Yes | Yes | Yes | Moderate |
| Raquel da Silva Ferreira | Species | Insect | Culex Spp. | Viral Prevalence | Yes | No | Yes | No | No | Yes | Yes | Yes | Yes | Moderate |
| Raquel da Silva Ferreira | Species | Insect | Psorophora albigenu | Viral Prevalence | Yes | No | Yes | No | No | Yes | Yes | Yes | Yes | Moderate |
| Carlos Borborema | Species | Insect | Culicoides paraensis | Seroprevalence | Unclear | Unclear | Yes | Yes | No | Yes | Yes | No | Yes | High |
| Carlos Borborema | Species | Insect | Culex quinquefasciatus | Seroprevalence | Unclear | Unclear | Yes | No | No | Yes | Yes | No | Yes | High |
| Carlos Borborema | Species | Insect | Culex quinquefasciatus | Seroprevalence | Unclear | Unclear | Yes | No | No | Yes | Yes | No | Yes | High |
| Carlos Borborema | Species | Insect | Culicoides paraensis | Seroprevalence | Unclear | Unclear | Yes | No | No | Yes | Yes | No | Yes | High |
| Carlos Borborema | Species | Insect | Culex quinquefasciatus | Seroprevalence | Unclear | Unclear | Yes | No | No | Yes | Yes | No | Yes | High |
| Pedro Fernando da Costa Vasconcelos | Age | Insect | Culicoides paraensis | Seroprevalence | Unclear | No | Yes | No | Yes | Yes | Yes | No | Yes | High |
| Pedro Fernando da Costa Vasconcelos | Age | Insect | Anopheles nuneztovari | Seroprevalence | Unclear | No | Yes | No | Yes | Yes | Yes | No | Yes | High |
| Pedro Fernando da Costa Vasconcelos | Age | Insect | Anopheles triannulatus | Seroprevalence | Unclear | No | Yes | No | Yes | Yes | Yes | No | Yes | High |

| | | | | | | | | | | | | | | |
|---|---|---|---|---|---|---|---|---|---|---|---|---|---|---|
| Pedro Fernando da Costa Vasconcelos | Age | Insect | Aedes scapularis | Seroprevalence | Unclear | No | Yes | No | Yes | Yes | Yes | No | Yes | High |
| Pedro Fernando da Costa Vasconcelos | Age | Insect | Psorophora cingulata | Seroprevalence | Unclear | No | Yes | No | Yes | Yes | Yes | No | Yes | High |
| Pedro Fernando da Costa Vasconcelos | Age | Insect | Psorophora ferox | Seroprevalence | Unclear | No | Yes | No | Yes | Yes | Yes | No | Yes | High |
| Pedro Fernando da Costa Vasconcelos | Age | Insect | Culex Spp. | Seroprevalence | Unclear | No | Yes | No | Yes | Yes | Yes | No | Yes | High |
| Pedro Fernando da Costa Vasconcelos | Age | Insect | Culex corniger | Seroprevalence | Unclear | No | Yes | No | Yes | Yes | Yes | No | Yes | High |
| Pedro Fernando da Costa Vasconcelos | Age | Insect | Culex coronator | Seroprevalence | Unclear | No | Yes | No | Yes | Yes | Yes | No | Yes | High |
| Pedro Fernando da Costa Vasconcelos | Age | Insect | Culex declarator | Seroprevalence | Unclear | No | Yes | No | Yes | Yes | Yes | No | Yes | High |
| Pedro Fernando da Costa Vasconcelos | Age | Insect | Culex quinquefasciatus | Seroprevalence | Unclear | No | Yes | No | Yes | Yes | Yes | No | Yes | High |
| Pedro Fernando da Costa | Age | Insect | Mansonia Sp. | Seroprevalence | Unclear | No | Yes | No | Yes | Yes | Yes | No | Yes | High |

| | | | | | | | | | | | | | | |
|---|---|---|---|---|---|---|---|---|---|---|---|---|---|---|
| Vasconcelos | | | | | | | | | | | | | | |
| Pedro Fernando da Costa Vasconcelos | Age | Insect | Limatus Sp | Seroprevalence | Unclear | No | Yes | No | Yes | Yes | Yes | No | Yes | High |
| Pedro Fernando da Costa Vasconcelos | Age | Insect | Sabethes glaucodaemon | Seroprevalence | Unclear | No | Yes | No | Yes | Yes | Yes | No | Yes | High |
| Pedro Fernando da Costa Vasconcelos | Age | Insect | Wyeomiya Sp. | Seroprevalence | Unclear | No | Yes | No | Yes | Yes | Yes | No | Yes | High |
| Pedro Fernando da Costa Vasconcelos | Age | Insect | Uranotaenia Sp | Seroprevalence | Unclear | No | Yes | No | Yes | Yes | Yes | No | Yes | High |
| Pedro Fernando da Costa Vasconcelos | Age | Insect | Culex Carrolia Sp | Seroprevalence | Unclear | No | Yes | No | Yes | Yes | Yes | No | Yes | High |
| Helver Dias | Species | Insect | Multiple | Viral Prevalence | Yes | No | Yes | Yes | Yes | Yes | Yes | Yes | Yes | Low |
| Helver Dias | Species | Insect | Wyeomiya Sp. | Viral Prevalence | Yes | No | Yes | Yes | Yes | Yes | Yes | Yes | Yes | Low |
| Helver Dias | Species | Insect | Aedes aegypti | Viral Prevalence | Yes | No | Yes | Yes | Yes | Yes | Yes | Yes | Yes | Low |
| Helver Dias | Species | Insect | Psorophora dimidiata | Viral Prevalence | Yes | No | Yes | No | Yes | Yes | Yes | Yes | Yes | Moderate |
| Helver Dias | Species | Insect | Culex nigripalpus | Viral Prevalence | Yes | No | Yes | Yes | Yes | Yes | Yes | Yes | Yes | Low |
| Helver Dias | Species | Insect | Psorophora albigenu | Viral Prevalence | Yes | No | Yes | Yes | Yes | Yes | Yes | Yes | Yes | Low |

| | | | | | | | | | | | | | | |
|---|---|---|---|---|---|---|---|---|---|---|---|---|---|---|
| Helver Dias | Species | Insect | Aedes scapularis | Viral Prevalence | Yes | No | Yes | No | Yes | Yes | Yes | Yes | Yes | Moderate |
| Helver Dias | Species | Insect | Aedes albopictus | Viral Prevalence | Yes | No | Yes | No | Yes | Yes | Yes | Yes | Yes | Moderate |
| Helver Dias | Species | Insect | Psorophora Spp. | Viral Prevalence | Yes | No | Yes | No | Yes | Yes | Yes | Yes | Yes | Moderate |
| Helver Dias | Species | Insect | Culex quinquefasciatus | Viral Prevalence | Yes | No | Yes | No | Yes | Yes | Yes | Yes | Yes | Moderate |
| Helver Dias | Species | Insect | Aedes Spp. | Viral Prevalence | Yes | No | Yes | No | Yes | Yes | Yes | Yes | Yes | Moderate |
| Helver Dias | Species | Insect | Haemagogus janthinomys | Viral Prevalence | Yes | No | Yes | No | Yes | Yes | Yes | Yes | Yes | Moderate |
| Helver Dias | Species | Insect | Psorophora cingulata | Viral Prevalence | Yes | No | Yes | No | Yes | Yes | Yes | Yes | Yes | Moderate |
| Helver Dias | Species | Insect | Psorophora cilipes | Viral Prevalence | Yes | No | Yes | No | Yes | Yes | Yes | Yes | Yes | Moderate |
| Helver Dias | Species | Insect | Mansonia Sp. | Viral Prevalence | Yes | No | Yes | No | Yes | Yes | Yes | Yes | Yes | Moderate |
| Helver Dias | Species | Insect | Psorophora lanei | Viral Prevalence | Yes | No | Yes | No | Yes | Yes | Yes | Yes | Yes | Moderate |
| Helver Dias | Species | Insect | Haemagogus leucocelaenus | Viral Prevalence | Yes | No | Yes | No | Yes | Yes | Yes | Yes | Yes | Moderate |
| Helver Dias | Species | Insect | Sabethes Spp. | Viral Prevalence | Yes | No | Yes | No | Yes | Yes | Yes | Yes | Yes | Moderate |
| Helver Dias | Species | Insect | Haemagogus Sp. | Viral Prevalence | Yes | No | Yes | No | Yes | Yes | Yes | Yes | Yes | Moderate |
| Helver Dias | Species | Insect | Culex Spp. | Viral Prevalence | Yes | No | Yes | Yes | Yes | Yes | Yes | Yes | Yes | Low |
| Helver Dias | Species | Insect | Wyeomiya Sp. | Viral Prevalence | Yes | No | Yes | No | Yes | Yes | Yes | Yes | Yes | Moderate |
| Helver Dias | Species | Insect | Aedes aegypti | Viral Prevalence | Yes | No | Yes | No | Yes | Yes | Yes | Yes | Yes | Moderate |

| | | | | | | | | | | | | | | |
|---|---|---|---|---|---|---|---|---|---|---|---|---|---|---|
| Helver Dias | Species | Insect | Psorophora dimidiata | Viral Prevalence | Yes | No | Yes | Yes | Yes | Yes | Yes | Yes | Yes | Low |
| Helver Dias | Species | Insect | Psorophora albigenu | Viral Prevalence | Yes | No | Yes | No | Yes | Yes | Yes | Yes | Yes | Moderate |
| Helver Dias | Species | Insect | Aedes scapularis | Viral Prevalence | Yes | No | Yes | No | Yes | Yes | Yes | Yes | Yes | Moderate |
| Helver Dias | Species | Insect | Aedes albopictus | Viral Prevalence | Yes | No | Yes | No | Yes | Yes | Yes | Yes | Yes | Moderate |
| Helver Dias | Species | Insect | Psorophora Spp. | Viral Prevalence | Yes | No | Yes | No | Yes | Yes | Yes | Yes | Yes | Moderate |
| Helver Dias | Species | Insect | Aedes Spp. | Viral Prevalence | Yes | No | Yes | No | Yes | Yes | Yes | Yes | Yes | Moderate |
| Helver Dias | Species | Insect | Haemagogus janthinomys | Viral Prevalence | Yes | No | Yes | No | Yes | Yes | Yes | Yes | Yes | Moderate |
| Helver Dias | Species | Insect | Psorophora cingulata | Viral Prevalence | Yes | No | Yes | No | Yes | Yes | Yes | Yes | Yes | Moderate |
| Helver Dias | Species | Insect | Psorophora cilipes | Viral Prevalence | Yes | No | Yes | No | Yes | Yes | Yes | Yes | Yes | Moderate |
| Helver Dias | Species | Insect | Mansonia Sp. | Viral Prevalence | Yes | No | Yes | No | Yes | Yes | Yes | Yes | Yes | Moderate |
| Helver Dias | Species | Insect | Psorophora lanei | Viral Prevalence | Yes | No | Yes | No | Yes | Yes | Yes | Yes | Yes | Moderate |
| Helver Dias | Species | Insect | Haemagogus leucocelaenus | Viral Prevalence | Yes | No | Yes | No | Yes | Yes | Yes | Yes | Yes | Moderate |
| Helver Dias | Species | Insect | Sabethes Spp. | Viral Prevalence | Yes | No | Yes | No | Yes | Yes | Yes | Yes | Yes | Moderate |
| Helver Dias | Species | Insect | Culex Spp. | Viral Prevalence | Yes | No | Yes | Yes | Yes | Yes | Yes | Yes | Yes | Low |
| Diego Michel Fernandes da Silva | Overall | Insect | Aedes aegypti | Viral Prevalence | Unclear | No | No | Yes | No | Yes | Yes | No | Yes | High |

| | | | | | | | | | | | | | | |
|---|---|---|---|---|---|---|---|---|---|---|---|---|---|---|
| F. P. Pinheiro | Species | Non-human animal | Multiple | Seroprevalence | Yes | Unclear | No | Yes | No | Yes | Yes | No | Yes | Moderate |
| F. P. Pinheiro | Species | Non-human animal | Multiple | Seroprevalence | Yes | Unclear | No | No | No | Yes | Yes | No | Yes | High |
| F. P. Pinheiro | Species | Non-human animal | Multiple | Seroprevalence | Yes | Unclear | No | Yes | No | Yes | Yes | No | Yes | Moderate |
| F. P. Pinheiro | Species | Non-human animal | Multiple | Seroprevalence | Yes | Unclear | No | Yes | No | Yes | Yes | No | Yes | Moderate |
| Paulo Mira Batista | Gender | Non-human animal | Non-human primates | Seroprevalence | Yes | No | Yes | No | Yes | Yes | Yes | Yes | Yes | Moderate |
| Paulo Mira Batista | Gender | Non-human animal | Non-human primates | Seroprevalence | Yes | No | Yes | No | Yes | Yes | Yes | Yes | Yes | Moderate |
| Paulo Mira Batista | Age | Non-human animal | Non-human primates | Seroprevalence | Yes | No | Yes | No | Yes | Yes | Yes | Yes | Yes | Moderate |
| Paulo Mira Batista | Age | Non-human animal | Non-human primates | Seroprevalence | Yes | No | Yes | No | Yes | Yes | Yes | Yes | Yes | Moderate |
| Paulo Mira Batista | Geography | Non-human animal | Non-human primates | Seroprevalence | Yes | No | Yes | No | Yes | Yes | Yes | Yes | Yes | Moderate |
| Paulo Mira Batista | Geography | Non-human animal | Non-human primates | Seroprevalence | Yes | No | Yes | No | Yes | Yes | Yes | Yes | Yes | Moderate |
| Paulo Mira Batista | Geography | Non-human animal | Non-human primates | Seroprevalence | Yes | No | Yes | No | Yes | Yes | Yes | Yes | Yes | Moderate |
| Paulo Mira Batista | Geography | Non-human animal | Non-human primates | Seroprevalence | Yes | No | Yes | No | Yes | Yes | Yes | Yes | Yes | Moderate |
| Paulo Mira Batista | Geography | Non-human animal | Non-human primates | Seroprevalence | Yes | No | Yes | No | Yes | Yes | Yes | Yes | Yes | Moderate |
| Helver Gonçalves Dias | Species | Non-human animal | Bos indicus/taurus | Seroprevalence | Yes | Yes | Yes | No | Yes | Yes | Yes | Yes | Yes | Moderate |

| | | | | | | | | | | | | | | |
|---|---|---|---|---|---|---|---|---|---|---|---|---|---|---|
| Helver Gonçalves Dias | Species | Non-human animal | Equus ferus caballus | Seroprevalence | Yes | Yes | Yes | No | Yes | Yes | Yes | Yes | Yes | Moderate |
| Helver Gonçalves Dias | Species | Non-human animal | Canis lupus familiaris | Seroprevalence | Yes | Yes | Yes | No | Yes | Yes | Yes | Yes | Yes | Moderate |
| Michael J. Turell | Overall | Non-human animal | Aotus nancymaae | Seroprevalence | No | Unclear | Yes | No | No | Yes | Yes | No | Yes | High |
| Plautino O. Laroque | Species | Non-human animal | Cebus flavius | Seroprevalence | Yes | Yes | Yes | No | No | Yes | Yes | Yes | Yes | Moderate |
| Plautino O. Laroque | Species | Non-human animal | Cebus libidinosus | Seroprevalence | Yes | Yes | Yes | Yes | Yes | Yes | Yes | Yes | Yes | Low |
| Alex Pauvolid-Correa | Species | Non-human animal | Equine species | Seroprevalence | Yes | Unclear | No | Yes | No | Yes | Yes | No | Yes | Moderate |
| Alex Pauvolid-Correa | Species | Non-human animal | Sheep species | Seroprevalence | Yes | Unclear | No | Yes | No | Yes | Yes | No | Yes | Moderate |
| Alex Pauvolid-Correa | Species | Non-human animal | Caiman species | Seroprevalence | Yes | Unclear | No | No | No | Yes | Yes | No | Yes | High |
| Helver Dias | Species | Non-human animal | Bos indicus/taurus | Viral Prevalence | Yes | No | Yes | No | Yes | Yes | Yes | Yes | Yes | Moderate |
| Helver Dias | Species | Non-human animal | Canis lupus familiaris | Viral Prevalence | Yes | No | Yes | No | Yes | Yes | Yes | Yes | Yes | Moderate |
| Helver Dias | Species | Non-human animal | Equus ferus caballus | Viral Prevalence | Yes | No | Yes | No | Yes | Yes | Yes | Yes | Yes | Moderate |
| Helver Dias | Species | Non-human animal | Felis silvestris catus | Viral Prevalence | Yes | No | Yes | No | Yes | Yes | Yes | Yes | Yes | Moderate |
| Helver Dias | Species | Non-human animal | Nasua nasua | Viral Prevalence | Yes | No | Yes | No | Yes | Yes | Yes | Yes | Yes | Moderate |
| Helver Dias | Species | Non-human animal | Didelphis albiventris | Viral Prevalence | Yes | No | Yes | No | Yes | Yes | Yes | Yes | Yes | Moderate |

| | | | | | | | | | | | | | | |
|---|---|---|---|---|---|---|---|---|---|---|---|---|---|---|
| Helver Dias | Species | Non-human animal | Mico melanurus | Viral Prevalence | Yes | No | Yes | No | Yes | Yes | Yes | Yes | Yes | Moderate |
| Helver Dias | Species | Non-human animal | Sapajus apella | Viral Prevalence | Yes | No | Yes | No | Yes | Yes | Yes | Yes | Yes | Moderate |
| Helver Dias | Species | Non-human animal | Didelphis aurita | Viral Prevalence | Yes | No | Yes | No | Yes | Yes | Yes | Yes | Yes | Moderate |
| Helver Dias | Species | Non-human animal | Aotus lemurinus | Viral Prevalence | Yes | No | Yes | No | Yes | Yes | Yes | Yes | Yes | Moderate |
| Helver Dias | Species | Non-human animal | Ateles marginatus | Viral Prevalence | Yes | No | Yes | No | Yes | Yes | Yes | Yes | Yes | Moderate |
| Helver Dias | Species | Non-human animal | Bos indicus /taurus | Viral Prevalence | Yes | No | Yes | Yes | Yes | Yes | Yes | Yes | Yes | Low |
| Helver Dias | Species | Non-human animal | Canis lupus familiaris | Viral Prevalence | Yes | No | Yes | Yes | Yes | Yes | Yes | Yes | Yes | Low |
| Helver Dias | Species | Non-human animal | Equus ferus caballus | Viral Prevalence | Yes | No | Yes | No | Yes | Yes | Yes | Yes | Yes | Moderate |
| Helver Dias | Species | Non-human animal | Felis silvestris catus | Viral Prevalence | Yes | No | Yes | No | Yes | Yes | Yes | Yes | Yes | Moderate |
| Helver Dias | Species | Non-human animal | Nasua nasua | Viral Prevalence | Yes | No | Yes | No | Yes | Yes | Yes | Yes | Yes | Moderate |
| Helver Dias | Species | Non-human animal | Didelphis albiventris | Viral Prevalence | Yes | No | Yes | No | Yes | Yes | Yes | Yes | Yes | Moderate |
| Helver Dias | Species | Non-human animal | Sapajus cay | Viral Prevalence | Yes | No | Yes | No | Yes | Yes | Yes | Yes | Yes | Moderate |
| Helver Dias | Species | Non-human animal | Callithrix jacchus | Viral Prevalence | Yes | No | Yes | No | Yes | Yes | Yes | Yes | Yes | Moderate |
| Helver Dias | Species | Non-human animal | Alouatta caraya | Viral Prevalence | Yes | No | Yes | No | Yes | Yes | Yes | Yes | Yes | Moderate |
| Paulo Mira Batista | Overall | Non-human animal | Multiple | Seroprevalence | No | Unclear | No | No | No | Yes | Yes | No | Yes | High |
| Scott Medlin | Species | Non-human animal | Bradypus variegatus | Seroprevalence | Unclear | Unclear | Yes | No | No | Yes | Yes | No | Yes | High |

| | | | | | | | | | | | | | | |
|---|---|---|---|---|---|---|---|---|---|---|---|---|---|---|
| Scott Medlin | Species | Non-human animal | Choloepus hoffmanni | Seroprevalence | Unclear | Unclear | Yes | No | No | Yes | Yes | No | Yes | High |
| Ana Cecilia Ribeiro Cruz | Species | Non-human animal | Phlegopsis n. pareensis | Seroprevalence | Yes | Unclear | Yes | No | No | Yes | Yes | Yes | Yes | Moderate |
| Ana Cecilia Ribeiro Cruz | Species | Non-human animal | Geotrygon montana | Seroprevalence | Yes | Unclear | Yes | No | No | Yes | Yes | Yes | Yes | Moderate |
| Ana Cecilia Ribeiro Cruz | Species | Non-human animal | Thamnophilus aethiops | Seroprevalence | Yes | Unclear | Yes | No | No | Yes | Yes | Yes | Yes | Moderate |
| Ana Cecilia Ribeiro Cruz | Species | Non-human animal | Conopophage roberti | Seroprevalence | Yes | Unclear | Yes | No | No | Yes | Yes | Yes | Yes | Moderate |
| Ana Cecilia Ribeiro Cruz | Species | Non-human animal | Schiffornis tudinus | Seroprevalence | Yes | Unclear | Yes | No | No | Yes | Yes | Yes | Yes | Moderate |
| Ana Cecilia Ribeiro Cruz | Species | Non-human animal | Xiphorhynchus ocellatus | Seroprevalence | Yes | Unclear | Yes | No | No | Yes | Yes | Yes | Yes | Moderate |
| Ana Cecilia Ribeiro Cruz | Species | Non-human animal | Myrmothera companisona | Seroprevalence | Yes | Unclear | Yes | No | No | Yes | Yes | Yes | Yes | Moderate |
| Ana Cecilia Ribeiro Cruz | Species | Non-human animal | Proechimis guianensis | Seroprevalence | Yes | Unclear | Yes | No | No | Yes | Yes | Yes | Yes | Moderate |
| Ana Cecilia Ribeiro Cruz | Species | Non-human animal | Didelphis marsupialis | Seroprevalence | Yes | Unclear | Yes | No | No | Yes | Yes | Yes | Yes | Moderate |
